# Repurposing cardiovascular disease prediction models for cancer

**DOI:** 10.64898/2026.02.03.26345370

**Authors:** Sam Quill, Aroon D. Hingorani, Nish Chaturvedi, Amand Floriaan Schmidt

## Abstract

**Background:** Population cancer screening detects the presence of early-stage disease rather than assessing future disease risk. We evaluated whether widely implemented cardiovascular disease (CVD) risk models can predict 10-year cancer risk and compared them with a less widely used cancer risk model (QCancer).

**Methods:** We evaluated four CVD prediction models: QRISK3, the Pooled Cohort Equations (PCE), SCORE2 and SCORE2-OP. All models were recalibrated using 20% of the UK Biobank (UKB) cohort and tested in the remainder, as well as in the Clinical Practice Research Datalink (CPRD). We gauged model performance using c-statistics for discrimination and evaluated the fidelity of calibration. We also identified the most influential risk factors in the QRISK3 model.

**Findings:** In the UKB test set, the c-statistics for incident CVD ranged from 0·71 to 0·74 (11,022 events). All CVD models achieved a c-statistic of 0·63 for any cancer (23,010 events) and showed CVD-equivalent discrimination for gastro-oesophageal, liver and biliary tree, laryngeal, renal tract, and lung cancers (c-statistic range: 0·70;0·81). Overall, the discrimination of the CVD models was comparable that of the QCancer models (median difference in c-statistic: -0·01 (95%CI -0·03;0·00). The recalibrated CVD models showed near-perfect calibration (median intercept 0·01, Q1;Q3 -0·05;0·03 and slope 1·00, Q1;Q3 0·93;1·15). Performance in CPRD (393,658 cancer events) was similar: the median c-statistic, calibration intercept, and slope were 0·01 (95%CI 0·00;0·02), 0·05 (95%CI 0·02;0·17), and 0·04 (95%CI 0·01;0·15) higher, respectively, in CPRD than in UKB. After age, smoking status and systolic blood pressure were the most influential predictors of cancer risk.

**Interpretation:** Widely implemented CVD prediction models perform similarly to the QCancer models in the prediction of incident cancers. They may be used to inform cancer prevention and guide risk-stratified monitoring. The recalibrated models are available through an API.

**Funding:** Health Data Research UK, British Heart Foundation and UK Research and Innovation.

## Background

Cancer and cardiovascular disease (CVD) are responsible for half of all deaths worldwide(1). The global health burden of these conditions is increasing, with cancer deaths expected to rise by approximately 75% by 2050(1). CVD and cancer share common risk factors, may co-occur, and are both potentially preventable via similar interventions. Addressing their modifiable risk factors is estimated to prevent over 75% CVD and 40% cancer diagnoses(2–4).

Despite these commonalities, early identification and intervention approaches differ. Risk models, such as QRISK3(5), are deployed to predict risk of CVD and as decision tools to commence medication for primary prevention. In contrast, for a limited range of cancers only (cervical, colorectal and breast), screening programmes use imaging or sampling to identify early disease. This targets people who benefit from cancer treatment but does not identify disease-free individuals who are at elevated risk of developing cancer in the future.

Clinical risk models that estimate future risk of cancer already exist (e.g. QCancer) but are not frequently used in practice(6). In contrast, CVD models are widely used. We therefore set out to explore whether CVD prediction models could be used to additionally identify people at risk of developing incident cancers.

We compared the performance of four CVD risk models in the prediction of the 11 cancer diagnoses against which the QCancer prediction models were developed and added four cancer diagnoses that are associated with high morbidity and mortality (laryngeal, brain, melanoma, liver and biliary tree cancers). We also evaluated whether the CVD models could predict the joint risk of people developing cancer and CVD, differentiating between cancer diagnoses before or after a CVD event.

## Methods

### Data resources

The UK Biobank (UKB) was used for the primary analysis, and the Clinical Practice Research Datalink (CPRD) Aurum database for external validation. The UKB recruited over half a million participants across the UK between 2006 and 2010, which we split into 20% training and 80% testing sets. CPRD provided longitudinal information on anonymised primary care records of 29,087,935 individuals in England from 1^st^ April 1997 to 29^th^ March 2021. These data resources have been described in detail previously(7,8).

Both UKB and CPRD were linked to Hospital Episode Statistics (HES), primary care data, and Office of National Statistics (ONS) death registry records. Linked data from the National Cancer Registration and Analysis Service (NCRAS) were available for the UKB cohort(8). We also used data collected at UKB registration, such as information on demographics, lifestyle, self-reported information including from nurse-led interview, and clinical data such as blood pressure, BMI and biochemistry data. To focus on clinically relevant subset of participants, we excluded participants under 40 or greater than 84 years of age at the start date of our study (1^st^ January 2011).

### Outcome definitions

We defined CVD as the composite of coronary heart disease, ischaemic stroke and sudden cardiac death. The any cancer endpoint included all cancers except non-melanomatous skin cancers. The specific cancer types we considered were: colorectal and anal, pancreatic, gastro-oesophageal (any stomach cancer or oesophageal cancer), liver and biliary tree (any liver cancer or cancer of the bile ducts), prostate, breast, ovarian, uterine, melanoma, brain and other intracranial cancers, oral, laryngeal, any blood cancer, renal tract (any kidney, ureter or bladder cancers), and lung cancers.

We considered the composite outcomes of any cancer, CVD or any cancer, and CVD and cancer. For participants who developed both CVD and cancer, we differentiated between CVD occurring after a cancer diagnosis, and CVD occurring prior to a cancer diagnosis. Finally, we included accidental injury as a negative control (see supplemental materials and Table S1 for the code lists used to define these outcomes). Prevalent conditions were defined as any diagnoses before or within one month of study start to accommodate recording delays(9).

### Implementation of risk prediction models

We calculated the predicted risk for all participants using the QRISK3, PCE, SCORE2, SCORE2-OP and QCancer models, applying the original model coefficients. We compared the predicted risk calculated by each model against the observed risk of each outcome at 10 years of follow up. The definition of the predictor variables for each model can be found in the supplemental materials and in Table S1-3. For composite outcomes without existing QCancer models (e.g., any type of cancer, CVD and cancer), we created a stacked QCancer model by taking the average of the logit-transformed predicted risks of all applicable cancer types (i.e., including prostate cancer for men and breast, ovarian and uterine cancers for women). The models were recalibrated (and independently evaluated) to improve an anticipated lack of calibration due to the difference between CVD incidence and that of individual cancers.

### Statistical analysis

The models were evaluated in terms of discrimination (c-statistic) and calibration (calibration intercept and calibration slope) including 95% confidence intervals (CIs). A c-statistic of 0.5 indicates the model is not able to discriminate between people who will and will not develop disease, whereas 1 represents perfect discrimination(10). Calibration measures how well the model’s predicted risk aligns with observed risk of experiencing the outcome being predicted. A calibration intercept of 0 and slope of 1 indicate perfect calibration(11).

To facilitate recalibration and unbiased evaluation of the prediction models, we first split the UKB cohort into 20% training and 80% testing sets, where the former was exclusively used to recalibrate the CVD and QCancer prediction models, and the testing set used to determine calibration and discrimination. Recalibration consisted of a rank-preserving (i.e., without affecting discrimination) transformation, deriving an outcome specific model update for the intercept and slope.

Permuted feature importance was performed on QRISK3, the most widely used CVD risk score in the UK. Each feature is randomly shuffled (50 times) in turn, with the resulting change in c-statistic representing the feature-specific contribution to model discrimination(16).

### Sensitivity and subgroup analyses

Our primary analysis focused on 10-year outcomes to align with the typical time frame used in CVD risk assessment for primary prevention. We additionally explored model performance over 1-, 2- and 5-years of follow up, and carried out a landmark analysis, removing any cancer event occurring within the first 3 years of available follow-up. This was to investigate the effects of removing sub-clinical or early onset cancers that may be less amenable to primary prevention interventions.

In the main analysis, we excluded participants with missing data for any of the predictor or outcome variables. As a sensitivity analysis, we re-ran the models after imputing missing data in the CPRD replication cohort using the *missRanger* package in R(13).

### Patient and public involvement and engagement

Patient representatives and members of the public helped to define the scope of outcomes to be included in the study and advised on communication of results to the wider public.

## Results

Data were available from 502,126 UKB and 4,810,089 CRPD participants. The mean age was 56·6 years (standard deviation (SD): 8·1) in UKB and 58·5 (SD: 12·1) in CPRD; 54·4% and 50% were female, respectively. The median follow-up for both UKB and CPRD was 10 years (UKB quartile(Q)1;Q3: 10;10, CPRD Q1;Q3: 6·1;10). Incident CVD developed in 26,821 (6·6%) and 244,280 (6·6%) respectively, incident cancer in 53,821 (11·7%) and 752,649 (17·8%) respectively, and both conditions in 3,954 (1·1%) and 53,131(1·6%) respectively; Supplemental Tables S4-5.

### Discrimination for incident CVD and cancers

The c-statistics for CVD at 10-years were 0·74 (95%CI 0·73;0·74) for QRISK3, 0·72 (95%CI 0·72;0·73) for PCE and SCORE2, and 0·71 (95%CI 0·71;0·72) for SCORE2-OP. The stacked QCancer model had a c-statistic of 0·65 (95%CI 0·64;0·65) for any cancer. This was higher than the CVD models, which all had a c-statistic of 0·63 (95%CI 0·63;0·64 for QRISK3 and SCORE2-OP, 95%CI 0·62;0·63 for PCE and SCORE2) for any cancer. All four CVD models had c-statistics of 0·70 or higher for the following individual cancers: gastro-oesophageal, liver and biliary tree, laryngeal, renal tract, any lung cancer, and lung cancer in smokers; Figure 1. Aside from the QCancer model’s markedly higher c-statistic for lung cancer, the discriminative performance was similar between the CVD and QCancer models (median difference: 0·01 95%CI 0·00;0·03).

**Figure 1.**
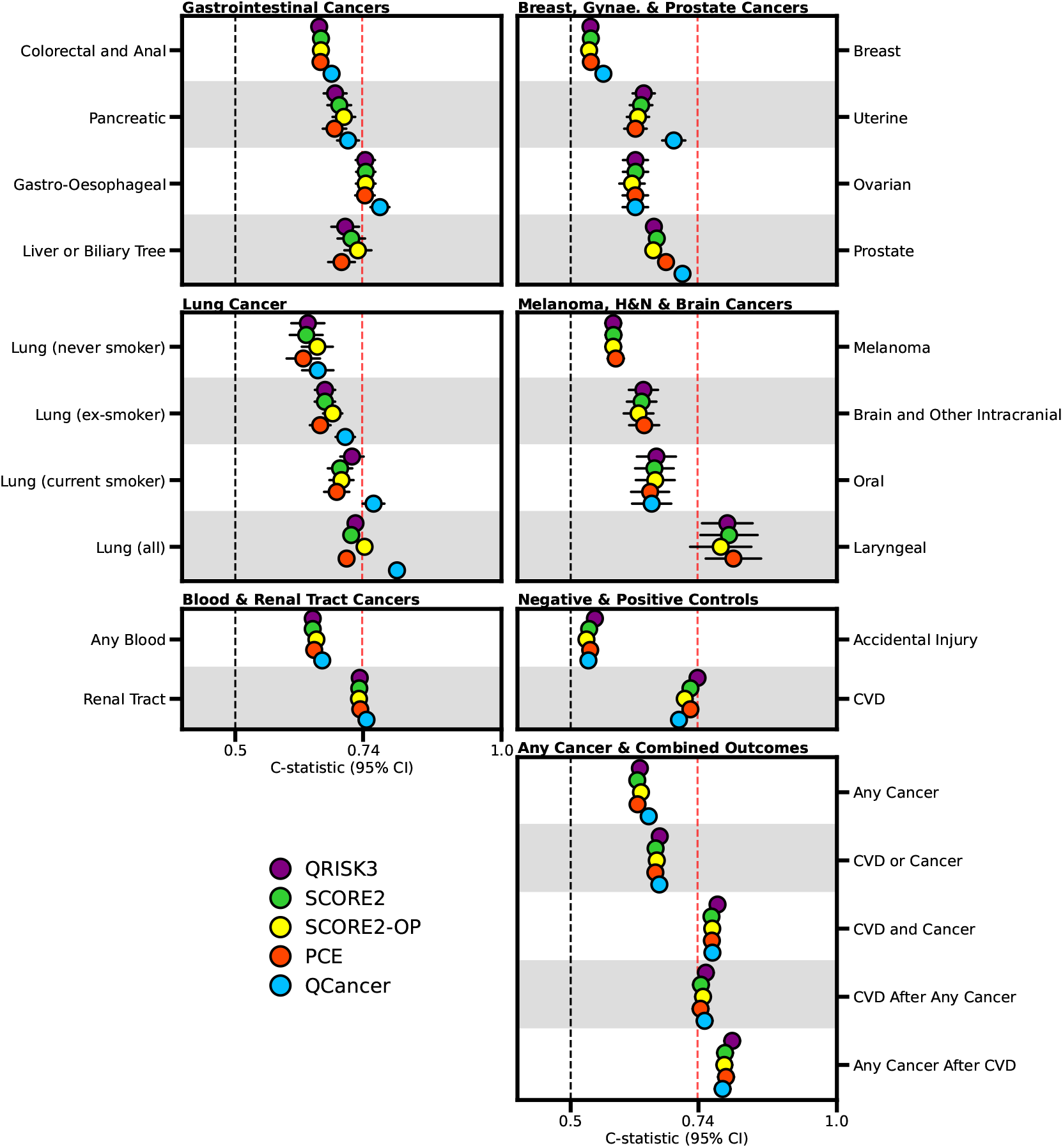
Discrimination of CVD and cancer risk prediction models for incident cancers, incident CVD, composite CVD and cancer outcomes, and accidental injury (negative control). The red, vertical dashed line represents the performance of QRISK3 for CVD at 10 years (c-statistic of 0·74, 95%CI :0·73; 0·74). The vertical dashed black line indicates the c-statistic value of 0·5. For each outcome, people with pre-existing disease (e.g. any cancer or any CVD) were excluded (Supplemental Tables S4-S5). For sex-specific cancers or cancers which are predominant in either males or females (incidence lower than 0·1%), performance was evaluated on the relevant sex-specific subgroup (Supplemental Tables S4-S5) Abbreviations: 95% CI, 95% confidence interval; CPRD, Clinical Practice Research Datalink; CVD, cardiovascular disease; Gynae. Gynaecological; H&N, head and neck; UKB, UK Biobank.

The CVD and QCancer models were able to identify people who developed both CVD and cancer (c-statistic range: 0·76 for SCORE2, 0·78 for QRISK3), and discrimination was higher still for cancer diagnoses following a CVD event (Figure 1 and Supplemental Table S6). The c-statistics for the negative control, accidental injury, ranged from 0·53 (95%CI 0·52;0·54) for SCORE2-OP to 0·55 (95%CI 0·54;0·55) for QRISK3.

### Calibration for incident CVD and cancers

Recalibrating the models improved the agreement between observed and predicted risk (evaluated in the 80% testing data) considerably. Overall, the calibration intercepts were close to 0 (median: 0·01, Q1;Q3: -0·05;0·03) and calibration slopes were close to 1 (median 1·00, Q1;Q3 0·92;1·15); Supplemental Table S6. There was no meaningful difference in calibration between the CVD and QCancer models (median difference: 0·00, 95%CI - 0·00;0·00 for the intercept; 0·03, 95%CI -0·03;0·12 for the slope); Figure 2, Supplemental Table S6.

**Figure 2.**
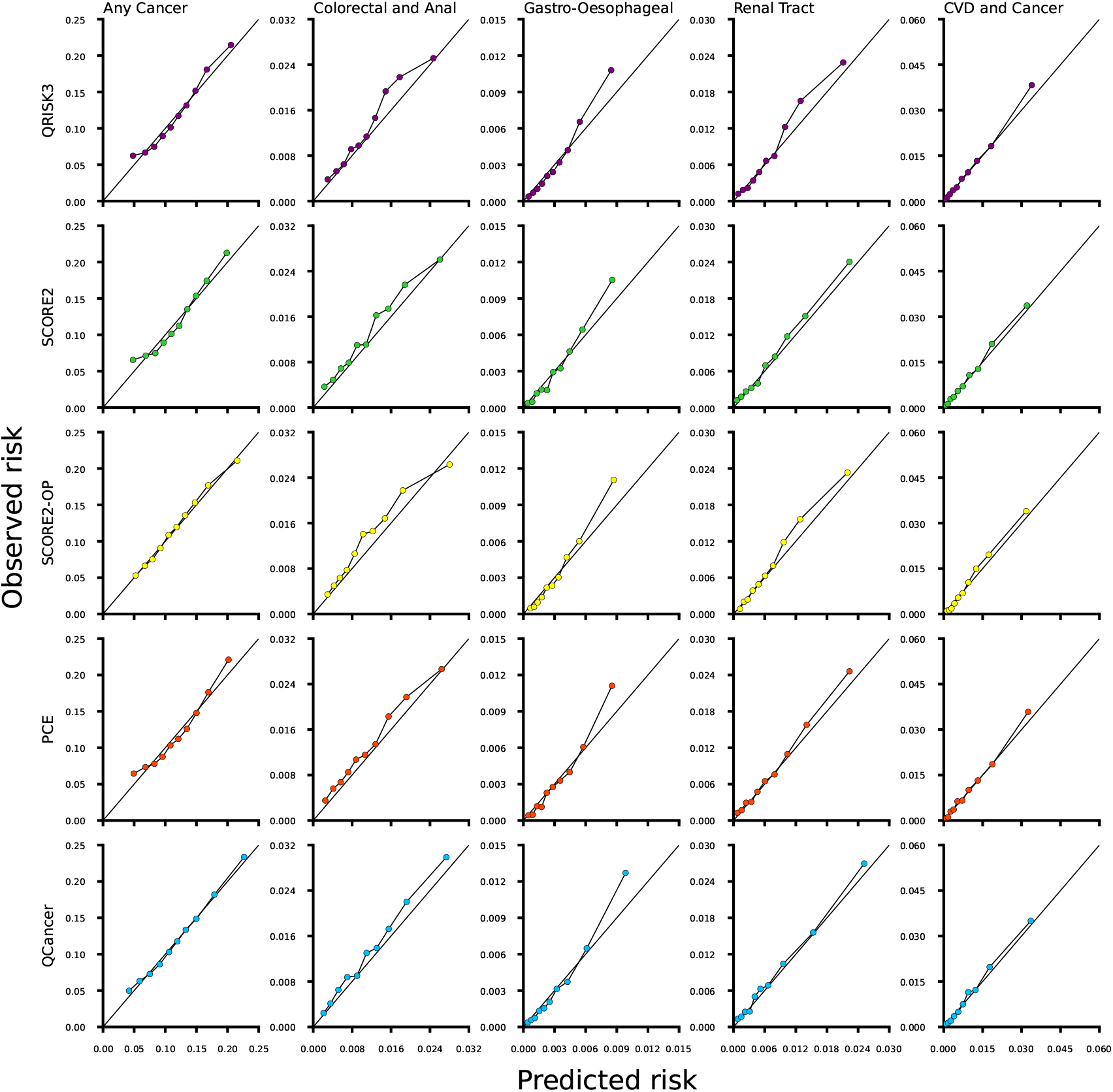
Calibration plots comparing the repurposed CVD risk models to QCancer risk models for different cancer outcomes in UK Biobank data after model recalibration. Calibration plots comparing predicted risk (x-axis) with observed risk (y-axis) between the four CVD risk models (QRISK3, PCE, SCORE2, SCORE2-OP) and QCancer model for five outcomes; any cancer, colorectal and anal cancer, gastro-oesophageal cancer, renal tract cancer, and the CVD and cancer endpoint. Each point represents a risk decile. Points on the solid diagonal line indicate perfect estimation of risk. Participants with missing predictor or outcome data were excluded from both cohorts. For each outcome, participants with pre-existing disease were excluded (Supplemental Tables S4-S5).

### External validation in CPRD

Model discrimination was similar across the UKB and CPRD. The median c-statistic for the CVD models was slightly higher in CPRD than in UKB (0.01, 95%CI 0.00;0.02). The c-statistics for all models were highly correlated between the two datasets; Spearman’s correlation (r) range 0·76 for PCE to 0·84 for QRISK3; Figure 3. Similarly, the difference in calibration between UKB and CPRD was small (median intercept difference: 0·05, 95%CI 0·01;0·17), median slope difference: 0·04 95%CI 0·01;0·15); Figure 3 and Table S6.

**Figure 3.**
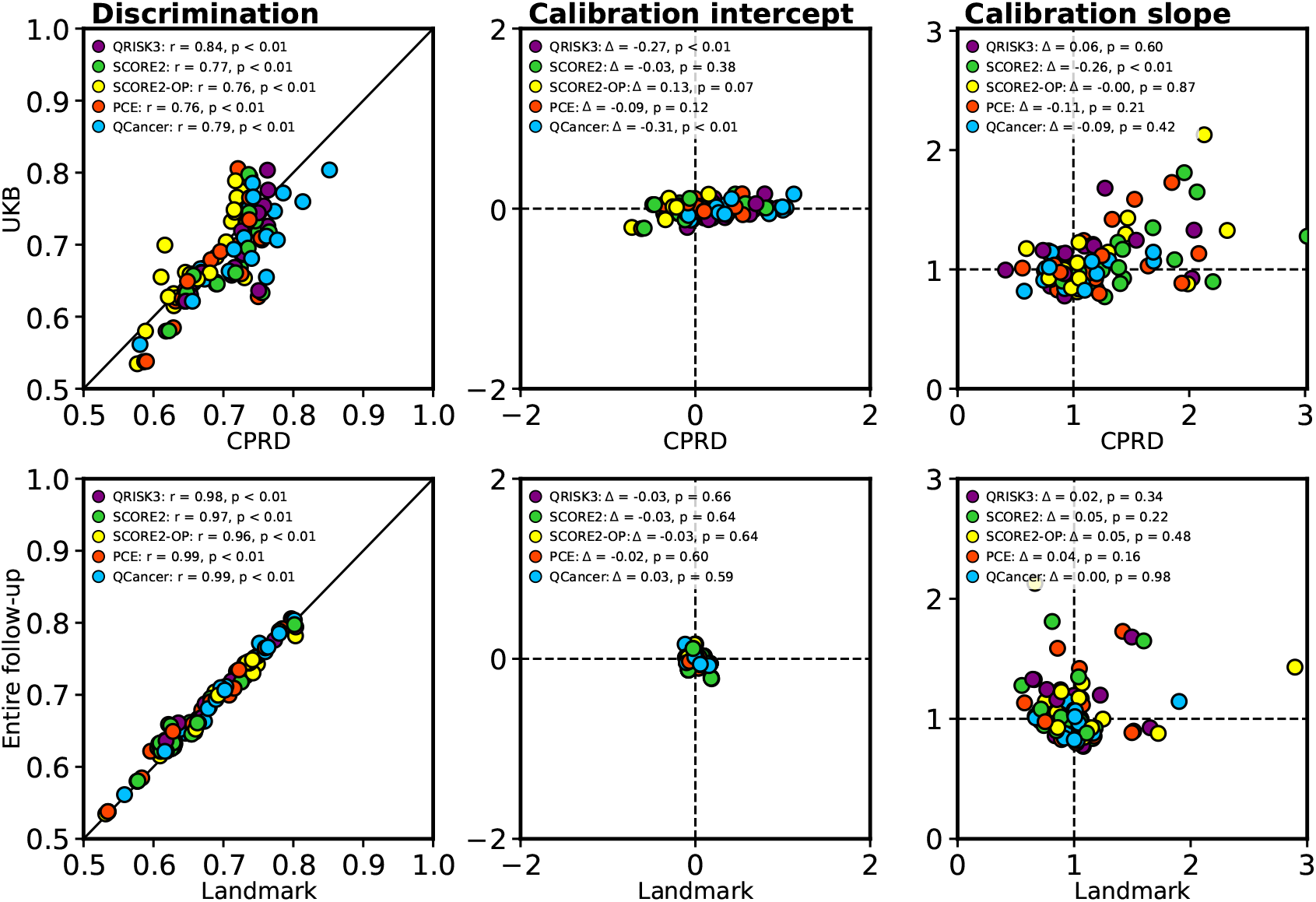
A comparison of the discrimination and calibration of each model between the UK Biobank and CPRD datasets, and for the landmark analysis on the UKB. Correlation between model performance metrics (discrimination, calibration intercept, and calibration slope) comparing UKB to CPRD (top row) and in the full UKB cohort to the landmark analysis excluding cancer diagnoses in the first 3 years of follow-up (bottom row). The correlation (r) was calculated using Spearman’s rank correlation test. The diagonal line in the discrimination plots (left column) indicates perfect correlation. The intersection of the dashed lines in the calibration intercept and slope plots (middle and right columns) indicates perfect agreement across datasets. The colours of the dots represent each of the models evaluated (see legend). Participants with missing predictor or outcome data were excluded from both cohorts. For each outcome, individuals with pre-existing disease were also excluded (see Supplemental Tables S4-S5). Δ indicates change between the dataset on the y-axis to the x-axis. Abbreviations: CPRD, Clinical Practice Research Datalink; UKB, UK Biobank.

### Sensitivity analyses

Discrimination did not meaningfully differ across the 1-, 2-, 5- and s10-year follow-up periods (Supplemental Figures S1 & S2). Similarly, the landmark analysis excluding cancers diagnoses in the first 3 years of follow-up did not meaningfully affect model performance (Figure 3 and Supplemental Table S7). Comparing the c-statistic results based on the complete-cases set of CPRD against the c-statistic using the imputed CPRD data showed reasonable agreement (correlation range between 0·78 and 0·83), with a median difference of 0·02 (95%CI - 0·02;0·03) (Supplemental Figure S3 and Table S6).

### Feature importance for predicting incident cancer

The permuted feature importance analysis indicated that smoking status was particularly important for nearly all cancers (aside from cancers of the prostate, uterus, ovaries, melanoma and smoking-stratified lung cancer), and specifically any laryngeal cancer and any lung cancer; Figure 4. Systolic blood pressure (SBP), total cholesterol to high-density lipoprotein cholesterol ratio, Townsend Deprivation Index, and family history of CVD showed a similar cross-cancer importance.

**Figure 4.**
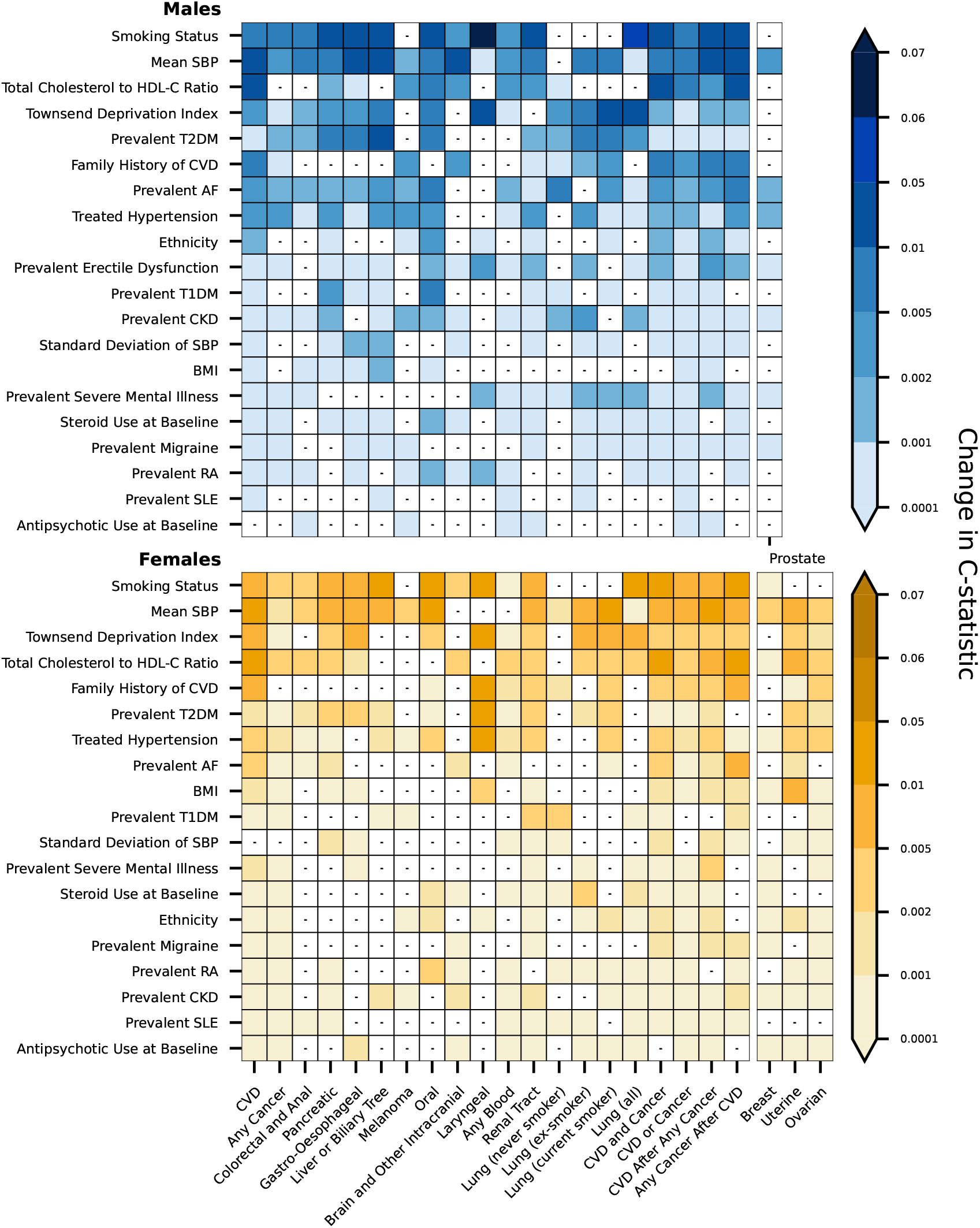
Feature importance in the QRISK3 model for cancer, cardiovascular disease and composite outcomes. The feature importance results for the QRISK3 model for each outcome, measured as the change in c-statistic when each variable is randomly permuted. The top panel (blue) shows results for male participants (including male-specific cancers) and the bottom panel (yellow) shows results for female participants (including female-specific cancers). Predictor variables (y-axis) are ordered by overall importance across all outcomes. The darker shading indicates greater importance (larger decrease in discrimination when variable is removed). The white tiles with a small, black horizontal line indicate which features did not contribute to the c-statistic in the QRISK3 model for that outcome. Abbreviations: AF, atrial fibrillation; BMI, body mass index; BP, blood pressure; CKD, chronic kidney disease; CVD, cardiovascular disease; HDL-C, high-density lipoprotein Cholesterol; RA, rheumatoid arthritis; SLE, systemic lupus erythematosus; T1DM, type 1 diabetes mellitus; T2DM, type 2 diabetes mellitus.

## Discussion

We successfully repurposed four widely used CVD prediction models (QRISK3, PCE and SCORE2 and SCORE2-OP) to additionally identify people at risk of developing cancers. The repurposed CVD models had comparable performance to the QCancer benchmark models for many common cancer types with established or emerging screening programmes, such as colorectal, breast, and prostate cancer. For lung cancer, the QCancer model had higher discrimination (c-statistic: 0.80 95%CI 0.79;0.81) than the repurposed CVD models overall (best c-statistic: 0.75 95%CI 0.74;0.76) for SCORE2-OP). However, among people with no history of smoking, the QCancer and repurposed CVD models performed similarly (c-statistic of 0.64 95%CI: 0.61;0.67 for QCancer and 0.63 95%CI: 0.059;0.66 for SCORE2-OP). The repurposed models had a discriminative performance similar to or better than that of CVD (c-statistic of 0·70 or higher) for several for several cancer types without established screening programmes, including renal tract, laryngeal, and gastro-oesophageal cancers, as well as cancers of the liver or biliary tree. The repurposed CVD models were able to identify people who developing both CVD and cancer (best c-statistic 0·77 95%CI 0·76;0·78 for QRISK3).

Although the original repurposed CVD models were generally poorly calibrated, recalibration improved this considerably in both UKB and on external validation in CPRD. In particular, the PCE model reached near-perfect calibration for many cancer types in both cohorts. The feature importance analysis showed that after age, smoking and SBP were the risk factors that contributed most to model performance in both men and women. These are well-established risk factors for many types of cancers, as well as for CVD, which provides a plausible explanation for why the repurposed CVD models also perform well when predicting cancer.

Disease prevention is a top priority for health systems worldwide. One of the core commitments in the UK’s *Fit for the Future* 10-Year Health Plan is to shift “from sickness to prevention” through early recognition and management of risk, long before disease develops(14). Despite overlapping risk factors and similar preventative interventions, primary prevention strategies for CVD and cancer have historically been implemented separately(15). Recognising the overlap between CVD and cancer risk, *Pursnani et al*. explored whether statin eligibility criteria overlapped with cancer risk(16), *Ogunmoroti et al*. determined the associations between Life’s Simple 7 lifestyle score and cancer incidence(17), and *Handy et al*. associated coronary artery calcifications scores with cancer risk(18). We sought to build on this work by developing formal cancer-specific risk prediction models, evaluating them in large sample size settings, and providing relevant external validation. We deliberately focused on repurposing models and data modalities that are currently available in most GP settings.

By repurposing CVD models already in active clinical use across Europe, the UK, and the US, our proposed approach could change the way cancer risk is managed in primary care, with limited additional resource demands for implementation. In the UK, the QRISK3 model is built into existing electronic health record systems e.g. in primary care and in the NHS Health Check, a national primary prevention programme for adults aged 40-74(19,20). This existing infrastructure for CVD risk assessment can readily be expanded to support cancer prevention and early detection initiatives, for example, generating prompts based on CVD risk scores that suggest interventions that reduce cancer risk or inviting high-risk individuals to join relevant cancer screening programmes.

Aligning cancer and CVD risk assessment using a single tool could have synergistic effects in preventing both conditions(15). Articulating elevated cancer risk alongside CVD risk would provide additional impetus for clinicians to encourage - and for patients to adopt - healthier lifestyles, alongside statin and antihypertensive therapy. Though medications are not currently used in cancer prevention, newer pharmacological agents, such as glucagon-like peptide-1 receptor agonists, may reduce risks of obesity-related cancers, which could lead to medicines having a more central role in primary prevention of cancer in the future(21,22).

While we sought to repurpose CVD models primarily to identify people at increased risk of developing novel cancers (rather than screen for subclinical disease), we acknowledge that at-risk individuals may require more focused monitoring. A possible avenue of future research might therefore be to explore the effectiveness of prioritised screening in people who are predicted to be at-risk of developing cancer. Both healthcare professionals and the general public are receptive to personalised, risk-based approaches to cancer screening(23). The repurposed CVD models could enable such risk-based cancer screening programmes to be rapidly adopted into clinical practice. In the UK, low-dose CT (LDCT) for lung cancer screening is offered to current or ex-smokers aged 55-74. Using our repurposed CVD models for recruitment would instead allow for lung cancer screening thresholds and intervals to be individualised based on predicted risk. Similar opportunities also exist around risk-based screening of both CVD and cancer, particularly given that LDCT may also be relevant for detecting early evidence of coronary artery calcification(24).

For cancers where population screening is not currently recommended due to limited test specificity (such as prostate-specific antigen testing for prostate cancer) or low disease incidence (e.g. renal and gastro-oesophageal cancers), targeting high-risk populations could make screening viable(46–49). Multi-cancer early detection tests, like the Galleri test®, would likely be more affordable if deployed in people with a relatively higher risk of developing cancer, rather than offering screening based on age alone(25).

There are several additional strengths of this study worth mentioning. Firstly, despite some differences in risk factor distributions and disease occurrence, we found comparably strong performances of the CVD models across both datasets for most outcomes. For example, average SBP was higher in UKB (138 mmHg; SD: 18·7) than in CPRD (132 mmHg; SD: 15·7), and fewer people developed cancer after 10 years of follow-up in UKB (53,821; 11·7%) compared to CPRD (752,649; 17·8%). Nonetheless, the median c-statistic was only 0·01 higher in CPRD compared to UKB (95%CI 0·00;0·02). This indicates the repurposed models are relatively robust to variation across different populations and likely reflects how they would perform in clinical practice.

Secondly, in our landmark analysis, we excluded all events within 3 years of baseline and demonstrated that the CVD models are able to identify cancers that develop *de novo*. This shows the repurposed CVD models can identify high-risk individuals at an early enough time, when risk reduction may prevent the development of cancer.

The following potential limitations require consideration. By restricting our analysis to adults aged 40-84 at study entry, we exclude young-onset cancers and cancers that develop at very advanced ages. However, young onset cancers typically have a distinct aetiology from cancers that develop in mid-to later life and are typically less amenable to primary prevention. Furthermore, some variables (e.g. BMI, cholesterol levels, smoking history) were only partially observed (particularly in the CPRD). However, repeating our analysis after imputing missing data in CPRD did not substantially affect the findings of the study. The limited impact of missing data on our results is also supported by the high degree of agreement between relatively simple models such as the PCE, which used nine predictors, and the more expansive QRISK3 model which requires information on over 20 variables.

## Conclusion

CVD prediction models can be repurposed to identify individuals at risk of cancer, as well as those at risk of both CVD and cancer. These repurposed, clinically used CVD models support deployment of targeted intervention for cancer prevention, as well as risk-based approaches to monitoring and cancer screening.

## Data Availability

Data are available from the UK Biobank and the Clinical Research Practice Datalink (CPRD).

## Availability of data and materials

The repurposed CVD models have been made available at: https://repurposed-cvd-risk-models.shinyapps.io/cvd_cancer_risk_app/

Data are available from the UK Biobank and the Clinical Practice Research Datalink.

## Author contributions

SQ, NC, AH and AFS designed the study. SQ performed the analyses and drafted the manuscript. NC, AH and AFS all provided critical input on the analysis, as well as on the drafted manuscript.

## Acknowledgments

This research has been conducted using the UK Biobank Resource under Application Number 12113. The authors are grateful to UK Biobank participants. This publication is part of the project “Computational medicine for cardiac disease” with file number 2025.027 of the research programme “Computing Time on National Computer Facilities” which is (partly) financed by the Dutch Research Council (NWO). The authors acknowledge the use of the UCL Myriad High Performance Computing Facility (Myriad@UCL), and associated support services, in the completion of this work.

## Code availability

All analyses were performed using R version 4.3.1. The figures were created using matplotlib, plot-misc(26), and Python version 3.1.1.

## Tables

**Table 1.** Baseline characteristics of participants in UK Biobank and CPRD cohorts.

| Variable | UKB Mean (SD) or N (%) | UKB Missing (%) | CPRD Mean (SD) or N (%) | CPRD Missing (%) |
| --- | --- | --- | --- | --- |
| **Overall sample size** | 502,126 |  | 4,810,089 |  |
| **Follow-up time in years (median, Q1/Q3)** | 10 (10; 10) |  | 10 (6·1; 10) |  |
| **Age (years)** | 56·6 (8·1) | 0 (0%) | 58·5 (12·1) | 0 (0%) |
| **Female sex (UKB), gender (CPRD)** | 273,158 (54·4%) | 0 (0%) | 2,404,320 (50%) | 24 (<0·1%) |
| **Mean systolic blood pressure (mmHg)** | 138 (18·7) | 1,318 (0·3%) | 132 (15·7) | 760,049 (15·8%) |
| **Total cholesterol (mmol/L)** | 5·7 (1·1) | 32,874 (6·5%) | 5·1 (1·1) | 1,744,818 (36·3%) |
| **HDL cholesterol (mmol/L)** | 1.4 (0.4) | 72,567 (14.5%) | 1.4 (0.4) | 2,037,512 (42.4%) |
| **BMI (kg/m<sup>2</sup>)** | 27.4 (4.8) | 3,328 (0.7%) | 27.3 (5.6) | 1,264,154 (26.3%) |
| **Personal history of:** |  |  |  |  |
| **Cardiovascular disease** | 33,768 (6.7%) |  | 602,809 (12.5%) |  |
| **Bowel cancer** | 2,860 (0.6%) |  | 30,007 (0.6%) |  |
| **Lung cancer** | 454 (0.1%) |  | 8,917 (0.2%) |  |
| **Breast cancer** | 11,449 (2.3%) |  | 72,016 (1.5%) |  |
| **Prostate cancer** | 3,570 (0.7%) |  | 43,889 (0.9%) |  |
| **Family history of:** |  |  |  |  |
| **Cardiovascular disease** | 215,137 (42.8%) | 10,746 (2.1%) | 1,062,660 (22.1%) | 0 (0%) |
| **Bowel cancer** | 54,590 (10.9%) | 91,262 (18.2%) | 58,262 (1.2%) | 0 (0%) |
| **Lung cancer** | 62,172 (12.4%) | 88,170 (17.6%) | 20,942 (0.4%) | 0 (0%) |
| **Breast cancer** | 52,464 (10.4%) | 91,759 (18.3%) | 79,216 (1.6%) | 0 (0%) |
| **Prostate cancer** | 38,760 (7.7%) | 95,388 (19%) | 4,174 (0.1%) | 0 (0%) |
| **Ethnicity:** |  | 895 (0.2%) |  | 245,474 (5.1%) |
| **White or not stated** | 474,238 (94.4%) |  | 4,160,043 (86.5%) |  |
| **Indian** | 5,944 (1.2%) |  | 90,799 (1.9%) |  |
| **Pakistani** | 1,833 (0.4%) |  | 39,744 (0.8%) |  |
| **Bangladeshi** | 236 (<0.1%) |  | 14,491 (0.3%) |  |
| **Other Asian** | 2,687 (0.5%) |  | 49,846 (1%) |  |
| **Black Caribbean** | 4,509 (0.9%) |  | 36,491 (0.8%) |  |
| **Black African** | 3,390 (0.7%) |  | 68,997 (1.4%) |  |
| **Chinese** | 1,573 (0.3%) |  | 17,562 (0.4%) |  |
| **Other ethnic group** | 6,821 (1.4%) |  | 86,624 (1.8%) |  |
| **Smoking status:** |  | 19,745 (3.9%) |  | 588,137 (12.2%) |
| **Non-smoker** | 273,336 (54.4%) |  | 2,351,207 (48.9%) |  |
| **Ex-smoker** | 172,923 (34.4%) |  | 1,234,591 (25.7%) |  |
| **<10 cigarettes/day** | 7,207 (1.4%) |  | 200,791 (4.2%) |  |
| **10-19 cigarettes/day** | 15,420 (3.1%) |  | 255,872 (5.3%) |  |
| **>19 cigarettes/day** | 13,495 (2.7%) |  | 179,491 (3.7%) |  |

Baseline characteristics are presented as mean (standard deviation) for continuous variables and frequency (percentage) for categorical variables. The percentage of missing data is shown for each variable within each cohort. BMI, body mass index; HDL-C, high-density lipoprotein cholesterol; UKB, UK Biobank; CPRD, Clinical Practice Research Datalink.

## References

1. Naghavi M, Kyu HH, A B, Aalipour MA, Aalruz H, Ababneh HS, et al. Global burden of 292 causes of death in 204 countries and territories and 660 subnational locations, 1990–2023: a systematic analysis for the Global Burden of Disease Study 2023. The Lancet. 2025 Oct 18;406(10513):1811–72.

2. Stewart J, Manmathan G, Wilkinson P. Primary prevention of cardiovascular disease: A review of contemporary guidance and literature. JRSM Cardiovasc Dis. 2017 Jan 1;6:2048004016687211.

3. Islami F, Marlow EC, Thomson B, McCullough ML, Rumgay H, Gapstur SM, et al. Proportion and number of cancer cases and deaths attributable to potentially modifiable risk factors in the United States, 2019. CA: A Cancer Journal for Clinicians. 2024;74(5):405–32.

4. Fink H, Langselius O, Vignat J, Rumgay H, Rehm J, Martinez RX, et al. Global and regional cancer burden attributable to modifiable risk factors to inform prevention. Nat Med. 2026 Feb 3;1–10.

5. Hippisley-Cox J, Coupland C, Brindle P. Development and validation of QRISK3 risk prediction algorithms to estimate future risk of cardiovascular disease: prospective cohort study. BMJ. 2017 May 23;357:2099.

6. Price S, Spencer A, Medina-Lara A, Hamilton W. Availability and use of cancer decision-support tools: a cross-sectional survey of UK primary care. Br J Gen Pract. 2019 Jul 1;69(684):e437–43.

7. Wolf A, Dedman D, Campbell J, Booth H, Lunn D, Chapman J, et al. Data resource profile: Clinical Practice Research Datalink (CPRD) Aurum. Int J Epidemiol. 2019 Dec 1;48(6):1740–1740g.

8. Sudlow C, Gallacher J, Allen N, Beral V, Burton P, Danesh J, et al. UK Biobank: An Open Access Resource for Identifying the Causes of a Wide Range of Complex Diseases of Middle and Old Age. PLOS Medicine. 2015 Mar 31;12(3):e1001779.

9. Lewis JD, Bilker WB, Weinstein RB, Strom BL. The relationship between time since registration and measured incidence rates in the General Practice Research Database. Pharmacoepidemiology and Drug Safety. 2005;14(7):443–51.

10. Harrell FE, Califf RM, Pryor DB, Lee KL, Rosati RA. Evaluating the yield of medical tests. JAMA. 1982 May 14;247(18):2543–6.

11. Van Calster B, McLernon DJ, van Smeden M, Wynants L, Steyerberg EW. Calibration: the Achilles heel of predictive analytics. BMC Med. 2019 Dec 16;17:230.

12. Altmann A, Toloşi L, Sander O, Lengauer T. Permutation importance: a corrected feature importance measure. Bioinformatics. 2010 May 15;26(10):1340–7.

13. Mayer M. missRanger: Fast Imputation of Missing Values [Internet]. 2025 [cited 2025 Oct 9]. Available from: https://github.com/mayer79/missRanger

14. NHS. Fit for the future: 10 Year Health Plan for England [Internet]. UK Government; 2025 [cited 2026 Jan 27]. Available from: https://assets.publishing.service.gov.uk/media/6888a0b1a11f859994409147/fit-for-the-future-10-year-health-plan-for-england.pdf

15. Handy CE, Quispe R, Pinto X, Blaha MJ, Blumenthal RS, Michos ED, et al. Synergistic Opportunities in the Interplay Between Cancer Screening and Cardiovascular Disease Risk Assessment. Circulation. 2018 Aug 14;138(7):727–34.

16. Pursnani A, Massaro JM, D’Agostino RB, O’Donnell CJ, Hoffmann U. Guideline-Based Statin Eligibility, Cancer Events, and Noncardiovascular Mortality in the Framingham Heart Study. J Clin Oncol. 2017 Sep 1;35(25):2927–33.

17. Ogunmoroti O, Allen NB, Cushman M, Michos ED, Rundek T, Rana JS, et al. Association Between Life’s Simple 7 and Noncardiovascular Disease: The Multi-Ethnic Study of Atherosclerosis. J Am Heart Assoc. 2016 Oct 20;5(10):e003954.

18. Handy CE, Desai CS, Dardari ZA, Al-Mallah MH, Miedema MD, Ouyang P, et al. The Association of Coronary Artery Calcium With Noncardiovascular Disease: The Multi-Ethnic Study of Atherosclerosis. JACC Cardiovasc Imaging. 2016 May;9(5):568–76.

19. Williams E, Round T, Jones NR. Cardiovascular disease — risk assessment and reduction: NICE 2023 update for GPs. Br J Gen Pract. 2024 Nov 1;74(748):523–6.

20. NICE. Cardiovascular disease: risk assessment and reduction, including lipid modification [A] Evidence review for CVD risk assessment tools: primary prevention [Internet]. National Institute for Health and Care Excellence (NICE); 2023 [cited 2024 Jan 26]. Available from: https://www.nice.org.uk/guidance/ng238/evidence/a-cvd-risk-assessment-tools-primary-prevention-pdf-13253901661

21. Wang L, Xu R, Kaelber DC, Berger NA. Glucagon-Like Peptide 1 Receptor Agonists and 13 Obesity-Associated Cancers in Patients With Type 2 Diabetes. JAMA Netw Open. 2024 Jul 5;7(7):e2421305.

22. Mao X, Zhang X, Henry L, Cheung KS, Yuen MF, Cheung R, et al. Association between glucagon-like peptidase 1 receptor agonist and obesity-related cancer in overweight or obese patients with type 2 diabetes: a nationwide cohort study. J Natl Cancer Inst. 2025 Oct 1;117(10):2053–61.

23. Tan NQP, Nargund RS, Douglas EE, Lopez-Olivo MA, Resong PJ, Ishizawa S, et al. Acceptability and perceptions of personalised risk-based cancer screening among health-care professionals and the general public: a systematic review and meta-analysis. The Lancet Public Health. 2025 Feb 1;10(2):e85–96.

24. Behr CM, IJzerman MJ, Kip MMA, Groen HJM, Heuvelmans MA, van den Berge M, et al. Model-Based Cost-Utility Analysis of Combined Low-Dose Computed Tomography Screening for Lung Cancer, Chronic Obstructive Pulmonary Disease, and Cardiovascular Disease. JTO Clin Res Rep. 2025 Feb 19;6(5):100813.

25. Kim A, Cong Z, Jazieh AR, Church TR, Reichert H, Nicholson G, et al. Estimating the incremental population health impact of a multi-cancer early detection (MCED) test to complement existing screening among elevated risk populations with multiple cancer risk factors: a mathematical modeling study. BMC Health Services Research. 2024 Dec 18;24(1):1584.

26. Amand Floriaan S. Plot-misc [Internet]. 2024 [cited 2025 Oct 9]. Available from: https://schmidtaf.gitlab.io/plot-misc/index.html

