## Supplementary Materials for "Repurposing cardiovascular disease prediction models for cancer"

### *Supplementary Methods*

#### *Data resources:*

##### *UKB*

The UK Biobank (UKB) recruited over half a million participants from across the UK aged from 37-73 years old between 2006 and 2010. We extracted data collected at the UKB index study visit, included demographic, lifestyle, biometric, and clinical data such as blood pressure and cholesterol measurements.

We derived baseline variables for the CVD and QCancer models in the UKB using both the data collected during the index study visit, and the participants' linked electronic health records (EHR). The EHR data included records from secondary care (Hospital Episode Statistics (HES)), primary care (available for approximately half of participants), cancer registry data, and Office of National Statistics death registry records(1). Personal and family history of relevant conditions was obtained from self-reported questionnaires. Self-reported information on medical history, medication use, and relevant clinical procedures was extracted from nurse-led interviews.

##### *CPRD*

All individuals in the Clinical Practice Research Datalink (CPRD) Aurum who were registered with participating GP practices that use the EMIS Web® system between 1st April 1997 and 29th March 2021 (the first and last date of HES data linkage) were potentially eligible for inclusion in the replication analysis. Linked data from secondary care (HES) data and the ONS death registry data was available for the majority of participants.

*Participant selection and eligibility*

To align with the intended target population of the risk prediction models, all participants aged 40 years or older and under 85 were eligible for inclusion in both the UKB and CPRD. Participants who died or were lost to follow-up between the study start date and the start follow period were excluded from UKB (n=44). Similarly, we excluded individuals who did not contribute to any follow-up time in CPRD, i.e. they deregistered from the practice, died or their data was last collected before the beginning of follow up were excluded (n=67,768).

Some additional processing steps were applied to CPRD that were not required in the UKB analysis. Only individuals whose records met CPRD's data quality standards were considered eligible(2). We excluded patients who had been registered at the GP practice for less than 1 year prior to the study start date (1st January 2011) to allow time for medical histories of newly registered patients to be captured in the EHR(3). We excluded participants who were ineligible to have their HES data linked to their primary care records(1). Finally, we excluded the small number of participants who had reported their gender as neither male nor female (n=24, <0·0001%) as the CVD models were not developed to accommodate non-binary gender variables (see Supplementary Tables S4-S5). Supplementary figure S4 is an example flow chart outlining the number of participants excluded from the any cancer analysis at each step of the data processing in UKB. Supplementary figure S5 shows a similar flow chart for the CPRD complete case and imputed datasets (see Supplementary Tables S4-S5for more details).

When preparing the dataset for analysis of a specific outcome, participants with pre-existing conditions relevant to that outcome were excluded. For cardiovascular disease (CVD) outcomes, we excluded participants with any evidence of coronary heart disease, any type of stroke, or sudden cardiac death prior to study enrolment. This exclusion criterion was intentionally broader than the CVD outcome definition itself (incident coronary heart disease, ischaemic stroke, or sudden cardiac death) to ensure we excluded participants with related (or potentially misclassified) prior events, such as subarachnoid haemorrhage. To align with how the QRISK3 model is implemented, people taking lipid lowering medications were also excluded when predicting outcomes that contained CVD (CVD, and the composite endpoints of CVD and cancer (see Supplementary Tables S4-S5).

Only participants with non-missing data for all model variables were included in the primary analysis to enable the models to be evaluated consistently across different outcomes (Supplementary Tables S4-S5).

**Data processing, UKB:**

*Outcome definitions in UKB*

We used data from UKB nurse-led surveys and questionnaires, primary care, secondary care diagnoses and procedures, death registry, and the cancer registry to define the models' input variables. As the touchscreen questionnaires and nurse-led interviews on self-reported data (such as personal medical history, medication use, family history of illness) were only carried out at baseline, incident outcomes were identified exclusively using the linked EHR data sources. Where possible, we used code lists from the HDR UK Phenotype Library as a starting point(4). ICD-10 codes for disease outcomes were mapped to their corresponding ICD-9 codes, and both were used to identify relevant Read codes and OPCS Classification of Interventions and Procedures (OPCS-4) codes, if applicable. For outcomes without suitable code lists in the HDR UK Phenotype Library (e.g., any cancer), we used manually defined ICD-10 codes, which were then mapped to ICD-9, Read, and OPCS-4 codes. Additional OPCS-4 codes were added manually, where indicated. The mapping was performed using UKB mapping files (UKB Resource 592, "all_lkps_maps_v4.xlsx")(5). All code lists underwent manual review to remove ineligible codes prior to use.

We included CVD as a positive control and accidental injury as a negative control in each model that we tested. Accidental injury was selected as a negative control because we assumed that its incidence would be independent of the predictors used in the CVD models. We defined accidental injury using ICD-10 codes V01-X59, excluding falls (W00-W19). We did this because as we assumed the incidence of falls is likely to be associated with underlying health status or functional impairments, which could also be associated with the predictors in the CVD model(6)

We constructed composite outcome variables (Supplementary Table S8) for several reasons: to enable comparison with composite outcomes evaluated by other purpose-built cancer models, e.g. the QCancer 10 year risk prediction model for gastro-oesophageal cancer(7); to investigate the ability of the models to predict a larger group of cancers at anatomically adjacent sites (e.g. renal tract cancer; liver and biliary tract cancer); and to investigate model performance for identifying combined CVD and cancer outcomes (either condition, or both). We also constructed the prevalent severe mental illness as a predictor variable to be used in the QRISK3 model. This was defined of having a personal medical history of depression, bipolar disorder or schizophrenia.

We excluded non-melanomatous skin cancer (NMSC) from our composite endpoint of any cancer. The majority of these NMSCs tend to be indolent, treatable cancers with excellent long-term prognoses. Basal cell carcinoma rarely metastasise and both basal and squamous cell carcinomas have very low mortality rates compared to other malignancies. NMSC cancers also have a much higher incidence than other cancer types(8). Including them in the composite cancer outcome would mean the any cancer outcome would be disproportionately weighted towards these cancer types that generally have low mortality rates.

For composite outcomes, we removed prevalent conditions related to either outcome. For example when preparing the run the models to predict CVD and cancer, we excluded participants with prevalent coronary heart disease, sudden cardiac death, any stroke, or any cancer (excluding NMSC) at baseline.

*Predictor variables in UKB*

*Medication use*

The codes used to define the predictor variables in UKB and CPRD can be found in Supplementary Tables S2 and S3 for the medication codes. The QRISK3 and QCancer algorithms define baseline medication use as at least two prescriptions, with the most recent treatment being prescribed within 4 weeks of the start of the study. The UKB did not collect data on the frequency and timing of prescriptions. We therefore assumed that participants who reported current medication use at their baseline visit were actively receiving these treatments.

*Ethnicity*

Self-reported ethnicity (UKB field ID: 21000) was used to create the different ethnicity variables required by each model. The categories we constructed were made to align with the definitions used in the original CVD and QCancer models (See Supplementary Table S8. Differences in predictor variables for the implemented cardiovascular risk prediction models. For more details).

*Family history of illness*

When family history of illness was elicited in the UKB surveys, the age the illness occurred in the family member was not recorded. Therefore, we assumed that the family history of heart disease variables refer to heart disease occurring in a first degree family member under the age of 60.

*Smoking status*

Smoking status was categorised as “never”, “former”, or “current” smokers for the SCORE2, SCORE2-OP and PCE models. For QRISK3 and QCancer, we used the information on cigarette quantity to subcategorise current smokers into light (fewer than10 cigarettes per day), moderate (between 10 and 20 cigarettes per day), and heavy (over 20 cigarettes per day) smoking categories supplementary the data tables.

*Alcohol intake*

The alcohol use variable required by the QCancer model was calculated using UKB touchscreen questionnaire data on the frequency and quantity of six alcoholic beverage types (red wine, white wine, beer/cider, spirits, fortified wine, and other alcoholic drinks). Participants reported either weekly or monthly consumption; monthly values were converted to weekly equivalents by dividing by 4·35. Each beverage type was converted to UK alcohol units using standard conversions: wine (2 units per glass), beer (2·5 units per glass), spirits (1 unit per measure), fortified wine (1·25 units per glass), and alcopops (1·3 units per bottle). Total weekly units were summed across all beverage types and divided by seven to calculate average daily alcohol units. Participants who reported never drinking alcohol were assigned zero units. Average daily alcohol consumption was then categorised according to QCancer definitions: none (0 units/day), trivial (<1 unit/day), light (1-2 units/day), moderate (3-6 units/day), heavy (7-9 units/day), and very heavy (>9 units/day). Responses coded as 'do not know' or 'prefer not to answer' were treated as missing. The *Calculating Alcohol Units* NHS resource was used to guide mappings of different alcohol types to estimates units of alcohol per drink(9).

**Data processing, CPRD**

*Outcome definitions in CPRD*

We aimed to maintain consistent definitions of model variables and outcomes across the primary and replication analysis. We mapped the disease code lists from the primary analysis from ICD10, ICD9, OPCS-4 and Read codes to SNOMED-CT codes using mapping files from the NHS Technology Reference Update Distribution (TRUD) website. SNOMED-CT codes were then matched to CPRD Aurum medcode IDs using CPRD's medcode dictionary. For each outcome, we searched the medcode dictionary “term” column using keywords to retrieve any medcode IDs that may have been missed during the mapping process. All lists underwent manual review to remove irrelevant or incorrectly mapped codes. These curated medcode lists were used to identify disease occurrence in CPRD primary care records (observation table).

For CPRD's linked HES data, we used the same ICD10 and OPCS-4 codes from the UKB primary analysis to identify cases of disease from secondary care records.

The date that the condition was first documented in primary care was available in CPRD using the observation table. However, this was not directly available in the HES Admitted Patient Care (APC) *diagnosis* or *procedure* tables in CPRD. We therefore assumed that an ICD10 coded diagnosis that a patient received during a hospital episode was made on the first day of the episode. Similarly, we used the first day of the hospital episode as a proxy for the date of diagnosis for conditions identified by OPCS-4 codes.

The earliest documented evidence of an outcome based on primary care, secondary care, medication use or linked death register data was used to define an incident case of the condition. Like in the UKB analysis, we assumed that individuals with no documented personal history of illness had no prior history of that condition.

*Predictor variables in CPRD*

*Medication use*

Medication use at baseline was defined using relevant BNF chapters (see supplementary table S6) and subchapters mapped to CPRD prodcodes. These prodcodes were then used to identify prescriptions in the CPRD *drugissue* table. Text searching of the prodcode dictionary in CPRD was additionally used to find any remaining prodcodes that should be included. In order to be classified as actively using a medication at baseline, participants required at least two prescription records in the prodcode list for that medication, with the most recent prescription issued within 1 month of the study start date.

*Ethnicity*

Ethnicity in CPRD was defined using HES data as the primary source. When HES ethnicity was unavailable, we used data from the CPRD observation table. We used SNOMED-CT code lists adapted from *Pineda-Moncusí et al.* (10) to classify participants into 12 initial categories: White, Unknown, Other Asian, Black African, Bangladeshi, Indian, Pakistani, Chinese, Black Caribbean, Other, Other Black, and Mixed. For QRISK3 and QCancer models, we combined categories to create nine groups: White or not stated (combining White and Unknown), Indian, Pakistani, Bangladeshi, Other Asian, Black Caribbean, Black African, Chinese, and Other ethnic group (combining Other, Other Black, and Mixed). The PCE model requires only three ethnicity categories so we simplified them to White (White or not stated), African American/Black (Black Caribbean and Black African), and Other (all remaining groups).

*Deprivation*

Index of multiple deprivation (IMD) was used in CPRD instead of Townsend Deprivation Index for the QRISK3 model as the Townsend Deprivation Index was not available. IMD is a relative measure based on seven domains: income, employment, education, crime, housing, health, and living environment(11). Where patient-level IMD was missing (~3% of the cohort), we used practice-level IMD instead. We used the 2019 IMD measures, as these were the earliest available to us. We assumed that deprivation status remained stable between each patient's registration date and 2019.

*Family history of illness*

Any previous evidence of family history of CVD or the various cancer types in the observation table was assumed to represent a positive family history of that condition. Similar to the UKB analysis, we assumed that any family history of a condition in a first degree relative occurred before the age of 60. In CPRD, we assumed that no documented evidence of family history in a first-degree relative constituted a negative family history for that illness.

*Clinical measurements*

We retrieved all systolic blood pressure measurements recorded within 5 years before each participant's study start date, excluding implausible values (<70 mmHg or >270 mmHg). The measurement closest to the study start date was used as baseline systolic blood pressure. Standard deviation of systolic blood pressure was calculated using a minimum of 2 and maximum of 5 measurements within this 5-year window.

We retrieved the most recent measurements recorded within 5 years before the study start date. If height and weight were recorded more recently than BMI, we calculated BMI from these values; otherwise, we used the recorded BMI directly. Implausible values (<9 kg/m² or >92 kg/m²) were excluded. BMI was set to missing if no valid measurements were available within the 5-year window.

For both total cholesterol and HDL cholesterol, we retrieved measurements recorded within 5 years of the study start date and used the value closest to study entry. Implausible values were excluded: total cholesterol <1 mmol/L or >20 mmol/L, and HDL cholesterol <0·1 mmol/L or >5 mmol/L. The total cholesterol to HDL ratio was calculated by dividing total cholesterol by HDL cholesterol from the most recent valid measurements.

The code lists we used to define these clinical measurements were created by text searching CPRD's medcode dictionary for relevant terms and supplementing this using published lists from OpenCodelists(12).

*Smoking status and alcohol use*

Smoking and alcohol intake were defined using the same categories as the primary UKB analysis. However, only records within 5 years of study start date were used to define smoking status and alcohol consumption. Unlike in the UKB analysis where there was an specific option for participants to decline to report their alcohol use, we assumed that missing responses for alcohol intake reflected no alcohol consumption.

In both UKB and CPRD, in order meet the criteria for having treated hypertension, participants were required to have a diagnosis of hypertension and be on antihypertensive medication at baseline (See supplementary table S6).

We did not use a participant's baseline HbA1c level (greater than or equal to 48 mmol/mol) in either UKB or CPRD to define type 2 diabetes, as we assumed this could not adequately differentiate between type 1 and type 2 diabetes. However, elevated HbA1c was included in the definition of any diabetes (used in the PCE model), which encompasses type 1 and type 2 diabetes, as well as forms of diabetes gestational diabetes and maturity onset diabetes of the young (MODY). Similarly, non-insulin antidiabetic medications were used to define type 2 diabetes at baseline, but insulin use alone was considered insufficiently specific for type 1 diabetes. Both insulin and non-insulin antidiabetic medications were included in the definition of any diabetes variable at baseline.

*Differences in predictor models between models*

Although the CVD models use many similar risk factors as predictors, some differences exist between how the variables were defined in each model. Supplementary table 2 provides an overview of how these similar predictors vary by CVD models. Supplementary table S8 provides an overview of the predictor variables used for each CVD model included in the study. Supplemental table S9 shows the same for the QCancer models.

**Supplemental methods**

*Risk model output:*

The CVD and QCancer risk prediction models generate continuous probability estimates ranging from 0 to 1. The probability estimates represent an individual's predicted risk of developing the outcome during a specific follow up time (i.e. 1-, 2-, 5-, 10 years).

*Incidence of cancer before and after CVD:*

The frequencies of the different cancer types occurring before and after CVD were counted and Chi-square tests with Bonferroni correction were used to investigate whether cancers were more likely to precede or follow CVD in patients who developed both CVD and cancer. We also calculated Kaplan-Meier estimates for each cancer type. These survival estimates were compared with the observed frequencies of different cancer types occurring before and after CVD to investigate whether the varying distribution of cancer types relative to the timing of a CVD diagnosis might be explained by differences in survival rates among different cancer types.

*Effect of the length of follow-up on discrimination*

To assess whether prediction performance varied with follow-up duration, we fit a linear regression model with the follow-up time as the input variable and the c-statistics as the outcome variable for each combination of outcome and time points. An arbitrary cutoff of a change in c-statistic by 0·02 was defined as a meaningful change in c-statistic across follow-up time points.

*Calculating median differences between models and datasets*

We calculated the median difference in performance between the CVD and QCancer models using Wilcox tests, after excluding the positive and negative controls (CVD and accidental injury). Wilcox tests were also used to compare the median model performance between datasets, e.g. model results in UKB compared to the model results in CPRD.

*Landmark analysis*

For the landmark analysis, cancers that developed within 3 years of the start of follow up were excluded and the median difference in c-statistic, calibration intercept and calibration slope was calculated across the datasets.

The supplementary figures were created using the plot-misc(13) library using Python version 3.1.1. and ggplot2(14) using R version 4.3.1.

This study was not registered nor was a study protocol developed.

The UK Biobank study have appropriate ethical approval in place: "UK Biobank has approval from the North West Multi-centre Research Ethics Committee (MREC) as a Research Tissue Bank (RTB) approval. This approval means that researchers do not require separate ethical clearance and can operate under the RTB approval (there are certain exceptions to this which are set out in the Access Procedures, such as re-contact applications)."

***Supplementary Results***

*Descriptive results*

There were more current smokers in CPRD (634,154; 13·2%) than in UKB (36,117; 7·2%) but fewer reported consuming any alcohol daily (1,057,706; 22% in CPRD, compared to 298,184 59% in UKB).·Mean systolic blood pressure was higher in UKB (138mmHg; SD:18·7 compared to 132 mmHg; SD:15·7 in CPRD), as were the total cholesterol levels (5·7mmol/L; SD 1·1, compared to 5·1mmol/L; SD 1·1 in CPRD). All family history variables were more frequency documented as positive in the UKB compared to CPRD, for example 215,137 (42·8%) in UKB compared to 1,062,660 (22·1%) in CPRD had a positive for family history of CVD.

*Exploring sequence of CVD and cancer occurrence*

Regarding the UKB participants who developed both cancer and CVD, we found that lung, pancreatic, and brain cancers occurred significantly more frequently after CVD than before it. We also found that breast and colorectal cancers occurred significantly more often before CVD than after (Supplementary Figure S6). Other cancer types like prostate and melanoma appeared more frequently before CVD than after, but these differences are not statistically significant. Similarly, gastro-oesophageal cancers and laryngeal cancers occurred more commonly after CVD, but the differences were also not statistically significant.

Cancer types that more frequently preceded CVD diagnosis demonstrated longer survival times. Those that occurred more frequently after CVD had shorter survival times (Supplementary Figure S7).

*Effect of the length of follow-up on discrimination*

The distribution of cancer incidence was relatively uniform over the UKB study follow up period (Supplementary Figure S8). None of the CVD models showed variation greater than a 0·02 change in c-statistic over the follow up time points, except for the PCE model when predicting lung cancer for current smokers and all models when predicting melanoma (Supplementary Figure S1). However, none of these changes were found to be statistically significant (Supplementary Figure S2).

***Supplemental References***

1. Sudlow C, Gallacher J, Allen N, Beral V, Burton P, Danesh J, et al. UK Biobank: An Open Access Resource for Identifying the Causes of a Wide Range of Complex Diseases of Middle and Old Age. PLOS Medicine. 2015 Mar 31;12(3):e1001779.

2. Wolf A, Dedman D, Campbell J, Booth H, Lunn D, Chapman J, et al. Data resource profile: Clinical Practice Research Datalink (CPRD) Aurum. Int J Epidemiol. 2019 Dec 1;48(6):1740–1740g.

3. Lewis JD, Bilker WB, Weinstein RB, Strom BL. The relationship between time since registration and measured incidence rates in the General Practice Research Database. Pharmacoepidemiology and Drug Safety. 2005;14(7):443–51.

4. HDR UK. HDR UK- Phenotype Library [Internet]. 2024 [cited 2024 Jan 19]. Available from: http://phenotypes.healthdatagateway.org/

5.: Resource 592 [Internet]. [cited 2025 Aug 31]. Available from: https://biobank.ndph.ox.ac.uk/ukb/refer.cgi?id=592

6. Arnold BF, Ercumen A, Benjamin-Chung J, Colford JMJ. Brief Report: Negative Controls to Detect Selection Bias and Measurement Bias in Epidemiologic Studies. Epidemiology. 2016 Sep;27(5):637.

7. Hippisley-Cox J, Coupland C. Development and validation of risk prediction algorithms to estimate future risk of common cancers in men and women: prospective cohort study. BMJ Open. 2015 Mar;5(3):e007825.

8. Rogers HW, Weinstock MA, Feldman SR, Coldiron BM. Incidence Estimate of Nonmelanoma Skin Cancer (Keratinocyte Carcinomas) in the US Population, 2012. JAMA Dermatol. 2015 Oct 1;151(10):1081–6.

9. nhs.uk [Internet]. 2022 [cited 2025 Aug 31]. Alcohol units. Available from: https://www.nhs.uk/live-well/alcohol-advice/calculating-alcohol-units/

10. Pineda-Moncusí M, Allery F, Delmestri A, Bolton T, Nolan J, Thygesen JH, et al. Ethnicity data resource in population-wide health records: completeness, coverage and granularity of diversity. Sci Data. 2024 Feb 22;11(1):221.

11. Noble M, Wright G, Smith G, Dibben C. Measuring Multiple Deprivation at the Small-Area Level. Environ Plan A. 2006 Jan 1;38(1):169–85.

12. OpenCodelists [Internet]. [cited 2025 Aug 30]. Available from: https://www.opencodelists.org/

13. Amand Floriaan S. Plot-misc [Internet]. 2024 [cited 2025 Oct 9]. Available from: https://schmidtaf.gitlab.io/plot-misc/index.html

14. Wickham H. Create Elegant Data Visualisations Using the Grammar of Graphics [Internet]. 2016 [cited 2025 Oct 9]. Available from: https://ggplot2.tidyverse.org/

*TRIPOD checklist*

TRIPOD+AI

Version: 11-January-2024

| Section/Topic | Item | Development / evaluation[1] | Checklist item | Reported on page |
|---|---|---|---|---|
| **TITLE** | | | | |
| *Title* | 1 | D;E | Identify the study as developing or evaluating the performance of a multivariable prediction model, the target population, and the outcome to be predicted | 1 |
| **ABSTRACT** | | | | |
| *Abstract* | 2 | D;E | See TRIPOD+AI for Abstracts checklist | 1 |
| **INTRODUCTION** | | | | |
| *Background* | 3a | D;E | Explain the healthcare context (including whether diagnostic or prognostic) and rationale for developing or evaluating the prediction model, including references to existing models | 3 |
| | 3b | D;E | Describe the target population and the intended purpose of the prediction model in the context of the care pathway, including its intended users (e.g., healthcare professionals, patients, public) | 3 |
| | 3c | D;E | Describe any known health inequalities between sociodemographic groups | n/a |
| *Objectives* | 4 | D;E | Specify the study objectives, including whether the study describes the development or validation of a prediction model (or both) | 3 |
| **METHODS** | | | | |
| *Data* | 5a | D;E | Describe the sources of data separately for the development and evaluation datasets (e.g., randomised trial, cohort, routine care or registry data), the rationale for using these data, and representativeness of the data | 3-4 & supplement |
| | 5b | D;E | Specify the dates of the collected participant data, including start and end of participant accrual; and, if applicable, end of follow-up | 3-4 & supplement |
| *Participants* | 6a | D;E | Specify key elements of the study setting (e.g., primary care, secondary care, general population) including the number and location of centres | 3-4 & supplement |
| | 6b | D;E | Describe the eligibility criteria for study participants | 3-4 & supp. |
| | 6c | D;E | Give details of any treatments received, and how they were handled during model development or evaluation, if relevant | n/a |
| *Data preparation* | 7 | D;E | Describe any data pre-processing and quality checking, including whether this was similar across relevant sociodemographic groups | Supplement |
| *Outcome* | 8a | D;E | Clearly define the outcome that is being predicted and the time horizon, including how and when assessed, the rationale for choosing this outcome, and whether the method of outcome assessment is consistent across sociodemographic groups | 4 |
| | 8b | D;E | If outcome assessment requires subjective interpretation, describe the qualifications and demographic characteristics of the outcome assessors | n/a |
| | 8c | D;E | Report any actions to blind assessment of the outcome to be predicted | n/a |
| *Predictors* | 9a | D | Describe the choice of initial predictors (e.g., literature, previous models, all available predictors) and any pre-selection of predictors before model building | 4-5 & supplement |
| | 9b | D;E | Clearly define all predictors, including how and when they were measured (and any actions to blind assessment of predictors for the outcome and other predictors) | 4-5 & supplement |
| | 9c | D;E | If predictor measurement requires subjective interpretation, describe the qualifications and demographic characteristics of the predictor assessors | n/a |
| *Sample size* | 10 | D;E | Explain how the study size was arrived at (separately for development and evaluation), and justify that the study size was sufficient to answer the research question. Include details of any sample size calculation | 3-4 & supplement |
| *Missing data* | 11 | D;E | Describe how missing data were handled. Provide reasons for omitting any data | 5 & supp. |
| *Analytical methods* | 12a | D | Describe how the data were used (e.g., for development and evaluation of model performance) in the analysis, including whether the data were partitioned, considering any sample size requirements | 3-4 & supplement |
| | 12b | D | Depending on the type of model, describe how predictors were handled in the analyses (functional form, rescaling, transformation, or any standardisation). | n/a |
| | 12c | D | Specify the type of model, rationale[2], all model-building steps, including any hyperparameter tuning, and method for internal validation | 5 & supplement |
| | 12d | D;E | Describe if and how any heterogeneity in estimates of model parameter values and model performance was handled and quantified across clusters (e.g., hospitals, countries). See TRIPOD-Cluster for additional considerations[3] | n/a |
| | 12e | D;E | Specify all measures and plots used (and their rationale) to evaluate model performance (e.g., discrimination, calibration, clinical utility) and, if relevant, to compare multiple models | 5 |
| | 12f | E | Describe any model updating (e.g., recalibration) arising from the model evaluation, either overall or for particular sociodemographic groups or settings | 5 |
| | 12g | E | For model evaluation, describe how the model predictions were calculated (e.g., formula, code, object, application programming interface) | 5 & 10 |
| *Class imbalance* | 13 | D;E | If class imbalance methods were used, state why and how this was done, and any subsequent methods to recalibrate the model or the model predictions | n/a |
| *Fairness* | 14 | D;E | Describe any approaches that were used to address model fairness and their rationale | n/a |
| *Model output* | 15 | D | Specify the output of the prediction model (e.g., probabilities, classification). Provide details and rationale for any classification and how the thresholds were identified | Supplement |

[1] D=items relevant only to the development of a prediction model; E=items relating solely to the evaluation of a prediction model; D;E=items applicable to both the development and evaluation of a prediction model

[2] Separately for all model building approaches.

[3] TRIPOD-Cluster is a checklist of reporting recommendations for studies developing or validating models that explicitly account for clustering or explore heterogeneity in model performance (eg, at different hospitals or centres). Debray et al, BMJ 2023; 380: e071018 [DOI: 10.1136/bmj-2022-071018]

| | | | | |
|---|---|---|---|---|
| *Training versus evaluation* | 16 | D;E | Identify any differences between the development and evaluation data in healthcare setting, eligibility criteria, outcome, and predictors | **6** |
| *Ethical approval* | 17 | D;E | Name the institutional research board or ethics committee that approved the study and describe the participant-informed consent or the ethics committee waiver of informed consent | **Supplement** |
| **OPEN SCIENCE** | | | | |
| *Funding* | 18a | D;E | Give the source of funding and the role of the funders for the present study | **2 & 11** |
| *Conflicts of interest* | 18b | D;E | Declare any conflicts of interest and financial disclosures for all authors | **Attached declarations** |
| *Protocol* | 18c | D;E | Indicate where the study protocol can be accessed or state that a protocol was not prepared | **Supplement** |
| *Registration* | 18d | D;E | Provide registration information for the study, including register name and registration number, or state that the study was not registered | **n/a** |
| *Data sharing* | 18e | D;E | Provide details of the availability of the study data | **10** |
| *Code sharing* | 18f | D;E | Provide details of the availability of the analytical code[4] | **10** |
| **PATIENT & PUBLIC INVOLVEMENT** | | | | |
| *Patient & Public Involvement* | 19 | D;E | Provide details of any patient and public involvement during the design, conduct, reporting, interpretation, or dissemination of the study or state no involvement. | **6** |
| **RESULTS** | | | | |
| *Participants* | 20a | D;E | Describe the flow of participants through the study, including the number of participants with and without the outcome and, if applicable, a summary of the follow-up time. A diagram may be helpful. | **Supplement** |
| | 20b | D;E | Report the characteristics overall and, where applicable, for each data source or setting, including the key dates, key predictors (including demographics), treatments received, sample size, number of outcome events, follow-up time, and amount of missing data. A table may be helpful. Report any differences across key demographic groups. | **6 & supplement** |
| | 20c | E | For model evaluation, show a comparison with the development data of the distribution of important predictors (demographics, predictors, and outcome). | **Supplement** |
| *Model development* | 21 | D;E | Specify the number of participants and outcome events in each analysis (e.g., for model development, hyperparameter tuning, model evaluation) | **Supplement** |
| *Model specification* | 22 | D | Provide details of the full prediction model (e.g., formula, code, object, application programming interface) to allow predictions in new individuals and to enable third-party evaluation and implementation, including any restrictions to access or re-use (e.g., freely available, proprietary)[5] | **10** |
| *Model performance* | 23a | D;E | Report model performance estimates with confidence intervals, including for any key subgroups (e.g., sociodemographic). Consider plots to aid presentation. | **6-9** |
| | 23b | D;E | If examined, report results of any heterogeneity in model performance across clusters. See TRIPOD Cluster for additional details[3]. | **n/a** |
| *Model updating* | 24 | E | Report the results from any model updating, including the updated model and subsequent performance | **7 & supp.** |
| **DISCUSSION** | | | | |
| *Interpretation* | 25 | D;E | Give an overall interpretation of the main results, including issues of fairness in the context of the objectives and previous studies | **6 & 8** |
| *Limitations* | 26 | D;E | Discuss any limitations of the study (such as a non-representative sample, sample size, overfitting, missing data) and their effects on any biases, statistical uncertainty, and generalizability | **10** |
| *Usability of the model in the context of current care* | 27a | D | Describe how poor quality or unavailable input data (e.g., predictor values) should be assessed and handled when implementing the prediction model | **n/a** |
| | 27b | D | Specify whether users will be required to interact in the handling of the input data or use of the model, and what level of expertise is required of users | **n/a** |
| | 27c | D;E | Discuss any next steps for future research, with a specific view to applicability and generalizability of the model | **8** |

From: Collins GS, Moons KGM, Dhiman P, et al. *BMJ* 2024;385:e078378. doi:10.1136/bmj-2023-078378

[4] This relates to the analysis code, for example, any data cleaning, feature engineering, model building, evaluation.
[5] This relates to the code to implement the model to get estimates of risk for a new individual.

***Supplementary tables***

Supplementary tables S1-7 can be found as attached files. Supplementary tables S8-12 are below.

Supplementary table S8. definitions of composite outcomes and their constituent disease codes

| Composite outcome | Codes of the constituent outcomes |
|---|---|
| Gastro-oesophageal cancer | Gastric cancer & oesophageal cancer |
| Renal tract cancer | Kidney cancer, ureter cancer, & bladder cancer |
| Liver and biliary tree cancer | Liver cancer & biliary tree cancer |
| Cardiovascular disease (CVD) | Coronary heart disease, ischaemic stroke, sudden cardiac death |
| CVD and cancer | CVD & any cancer |
| CVD of cancer | CVD & any cancer |
| CVD after cancer | CVD & any cancer |
| Cancer after CVD | CVD & any cancer |

*This table indicates how the composite outcomes (leftmost column) were defined. Any codes present in the constituent outcomes (rightmost column) were included in the broader definition of the composite outcomes.*

Supplementary table S9 showing the input variables used the four CVD models evaluated in this study.

| Predictor | QRISK3 | PCE | SCORE2 | SCORE2-OP |
|---|---|---|---|---|
| Population in which the tool was derived | UK | US | European-wide | European-wide |
| Age | ✓ | ✓ | ✓ | ✓ |
| Sex | ✓ | ✓ | ✓ | ✓ |
| Smoking Status | ✓ | ✓ | ✓ | ✓ |
| Systolic blood pressure | ✓ | ✓ | ✓ | ✓ |
| Serum cholesterol levels | ✓ | ✓ | ✓ | ✓ |
| Area of residence | ✓ | | | |
| Risk region (country of residence) | | | ✓ | ✓ |
| Ethnicity | ✓ | ✓ | | |
| History of diabetes | ✓ | ✓ | | |
| Treated hypertension | ✓ | ✓ | | |
| Body mass index | ✓ | | | |
| Family history of CVD | ✓ | | | |
| Chronic kidney disease | ✓ | | | |
| Atrial fibrillation | ✓ | | | |
| Migraines | ✓ | | | |
| Rheumatoid arthritis | ✓ | | | |
| Systemic lupus erythematosus | ✓ | | | |
| Severe mental illness | ✓ | | | |
| Erectile dysfunction | ✓ | | | |
| Corticosteroid use | ✓ | | | |
| Using second generation antipsychotics | ✓ | | | |

*This table shows the predictor variable used in the four models included in this study. Note: CVD: cardiovascular diseas, PCE: Pooled Cohort Equations, SCORE2: Systematic Coronary Risk Evaluation 2; SCORE2-OP: Systematic Coronary Risk Evaluation 2 - Older Persons; UK: United Kingdom; US: United States; Family history of CVD: Family history of angina or heart attack in first degree relative under 60 years of age. BNF: British National Formulary*

Supplementary table S10: Input variables for the Qcancer 10 year risk models.

| Predictor | Uterine | Ovarian | Breast | Prostate | Lung | Blood | Renal | CRC | G-O | Panc | Oral |
|---|---|---|---|---|---|---|---|---|---|---|---|
| Age | ✓ | ✓ | ✓ | ✓ | ✓ | ✓ | ✓ | ✓ | ✓ | ✓ | ✓ |

| | | | | | | | | | | | |
|---|---|---|---|---|---|---|---|---|---|---|---|
| Sex | | | | | ✓ | ✓ | ✓ | ✓ | ✓ | ✓ | ✓ |
| Ethnicity | | | ✓ | ✓ | ✓ | | | ✓ | | | |
| Deprivation index | | | ✓ | ✓ | ✓ | | ✓ | ✓ | ✓ | ✓ | ✓ |
| Smoking status | ✓ | | | ✓ | ✓ | ✓ | ✓ | ✓ | ✓ | ✓ | ✓ |
| Alcohol intake | | | ✓ | | ✓ | | | ✓ | ✓ | | ✓ |
| Body mass index | ✓ | ✓ | ✓ | ✓ | ✓ | ✓ | ✓ | ✓ | ✓ | ✓ | ✓ |
| Taking OCP | | ✓ | ✓ | | | | | | | | |
| Taking oestrogen-containing HRT | | | ✓ | | | | | | | | |
| *Family history of:* | Uterine | Ovarian | Breast | Prostate | Lung | Blood | Renal | CRC | G-O | Panc | Oral |
| Gastro-intestinal cancer | | | | | | | | ✓ | | | |
| Lung cancer | | | | | ✓ | | | | | | |
| Blood cancer | | | | | | ✓ | | | | | |
| Breast cancer | | | ✓ | | | | | | | | |
| Ovarian cancer | | ✓ | | | | | | | | | |
| Prostate cancer | | | | ✓ | | | | | | | |
| *Personal history of:* | Uterine | Ovarian | Breast | Prostate | Lung | Blood | Renal | CRC | G-O | Panc | Oral |
| Colorectal cancer | ✓ | | | | ✓ | | ✓ | | | | ✓ |
| Gastro-oesophageal cancer | | | | | ✓ | | | | | | |
| Pancreatic cancer | | | | | | | | | ✓ | | |
| Oral cancer | | | | | ✓ | | | ✓ | ✓ | | |
| Lung cancer | | | ✓ | | | | ✓ | ✓ | ✓ | | ✓ |
| Blood cancer | | | ✓ | | ✓ | | ✓ | ✓ | ✓ | ✓ | ✓ |
| Bladder or kidney cancer | | | | | ✓ | ✓ | | | | ✓ | |
| Brain tumour | | | | | | ✓ | ✓ | | | | |
| Breast cancer | ✓ | ✓ | | | ✓ | | | ✓ | ✓ | ✓ | |
| Ovarian cancer | | | ✓ | | ✓ | ✓ | ✓ | ✓ | | | ✓ |
| Uterine cancer | | | | | ✓ | | ✓ | ✓ | | | |
| Cervical cancer | | ✓ | | | ✓ | | ✓ | ✓ | | | |
| Prostate cancer | | | | | | | ✓ | | | | |
| Type 1 diabetes | | | | ✓ | | ✓ | | | | | |
| Type 2 diabetes | ✓ | | | ✓ | | | ✓ | ✓ | ✓ | ✓ | |
| Peptic ulcer disease | | | | | | | | | ✓ | | |
| Barrett's oesophagus | | | | | | | | | ✓ | | |
| Chronic pancreatitis | | | | | | | | | | ✓ | |
| Ulcerative colitis | | | | | | | | ✓ | | | |
| Colonic polyps | | | | | | | | ✓ | | | |
| COPD | | | | | ✓ | | | | | | |
| Asthma | | | | | ✓ | | | | | | |
| Asbestos exposure | | | | | ✓ | | | | | | |
| Bipolar or schizophrenia | | | ✓ | ✓ | | | | | | | |
| Benign breast disease | | | ✓ | | | | | | | | |
| Polycystic ovaries | ✓ | | | | | | | | | | |
| Endometrial polyps or hyperplasia | ✓ | | | | | | | | | | |

*This table shows the predictor variables used in the QCancer model. The tick (✓) indicates that this variable is used as an input variable in that QCancer 10-year risk model CRC: colorectal cancer; G-O: gastro-oesophageal Cancer; Panc: pancreatic cancer; OCP: Oral contraceptive pill; HRT: hormone replacement therapy*

Supplementary Table S11. Differences in predictor variables for the implemented cardiovascular risk prediction models.

| Predictor | QRISK3 | PCE | SCORE-2 | SCORE-2 OP |
|---|---|---|---|---|
| Smoking Status | 5 levels: Non-smoker, ex-smoker, light smoker (<10 a day), moderate smoker (10–19 a day), or heavy smoker (20 or over a day). | Smoker, 2 levels: current, other. | Smoker, 2 levels: current, other. | Smoker, 2 levels: current, other. |
| Area of residence (deprivation index) | Townsend deprivation score (Area-level deprivation) | N/A | Risk region, 3 levels: low, moderate or high | Risk region, 3 levels: low, moderate or high |
| History of diabetes | Personal history of type 1 or type 2 diabetes mellitus. | Personal history of any type of diabetes mellitus (i.e. including, for example, gestational diabetes). | N/A | N/A |
| Ethnicity | 9 levels: White or not stated, Indian, Pakistani, Bangladeshi, Other Asian, Black Caribbean, Black African, Chinese, or "other ethnic group". | 3 levels: White, African American, Other. | N/A | N/A |

*Supplementary Table S11. Differences in definitions of predictor variable of the source studies deriving the implementing the cardiovascular risk prediction models. Note PCE: Pooled Cohort Equations, SCORE-2: Systematic Coronary Risk Evaluation 2; SCORE2-OP: Systematic Coronary Risk Evaluation 2 - Older Persons;) SD: Standard Deviation; HDL: high density lipoprotein. N/A: not applicable.*

Supplementary table S12: descriptive statistics of the imputed CPRD dataset.

| Variable | CPRD Mean (SD) or N (%) | CPRD Missing (%) |
|---|---|---|
| **Overall sample size** | 4,810,089 | |
| **Follow-up time in years (median, Q1/Q3)** | 10 (10; 10) | |
| **Age (years)** | 58·5 (12·1) | 0 (0%) |
| **Female Gender (CPRD)** | 2,404,320 (50%) | 0 (0%) |
| **Mean systolic blood pressure (mmHg)** | 132 (15·7) | 0 (0%) |
| **Total Cholesterol (mmol/L)** | 5·3 (1·1) | 0 (0%) |
| **HDL Cholesterol (mmol/L)** | 1·5 (0·5) | 0 (0%) |
| **BMI (kg/m$^2$)** | 27·1 (5·5) | 0 (0%) |
| **Personal History of:** | | |
| **Cardiovascular disease** | 602,809 (12·5%) | |
| **Bowel cancer** | 30,007 (0·6%) | |
| **Lung cancer** | 8,917 (0·2%) | |
| **Breast cancer** | 72,016 (1·5%) | |
| **Prostate cancer** | 43,889 (0·9%) | |
| **Family history of:** | | |
| **Cardiovascular disease** | 1,062,660 (22·1%) | 0 (0%) |
| **Bowel cancer** | 58,262 (1·2%) | 0 (0%) |
| **Lung cancer** | 20,942 (0·4%) | 0 (0%) |
| **Breast cancer** | 79,216 (1·6%) | 0 (0%) |
| **Prostate cancer** | 4,174 (0·1%) | 0 (0%) |
| **Ethnicity:** | | 0 (0%) |
| **White or not stated** | 4,379,362 (91%) | |
| **Indian** | 96,562 (2%) | |
| **Pakistani** | 42,371 (0·9%) | |
| **Bangladeshi** | 15,646 (0·3%) | |
| **Other Asian** | 52,786 (1·1%) | |
| **Black Caribbean** | 38,690 (0·8%) | |
| **Black African** | 73,705 (1·5%) | |
| **Chinese** | 18,691 (0·4%) | |
| **Other ethnic group** | 92,276 (1·9%) | |

| | | |
|---|---|---|
| **Smoking status:** | | 0 (0%) |
| **Non-smoker** | 2,654,032 (55·2%) | |
| **Ex-smoker** | 1,401,403 (29·1%) | |
| **<10 cigarettes/day** | 236,698 (4·9%) | |
| **10-19 cigarettes/day** | 303,642 (6·3%) | |
| **>19 cigarettes/day** | 214,314 (4·5%) | |

*Supplemental table S12: baseline characteristics for the imputed CPRD dataset are shown as mean (standard deviation) for continuous variables and frequency (percentage) for categorical variables. The percentage of missing data is shown for each variable within each cohort. BMI, body mass index; HDL-C, high-density lipoprotein cholesterol; CPRD, Clinical Practice Research Datalink.*

### *Supplementary figures*

#### Figure S1: change in c-statistic over the follow up time points

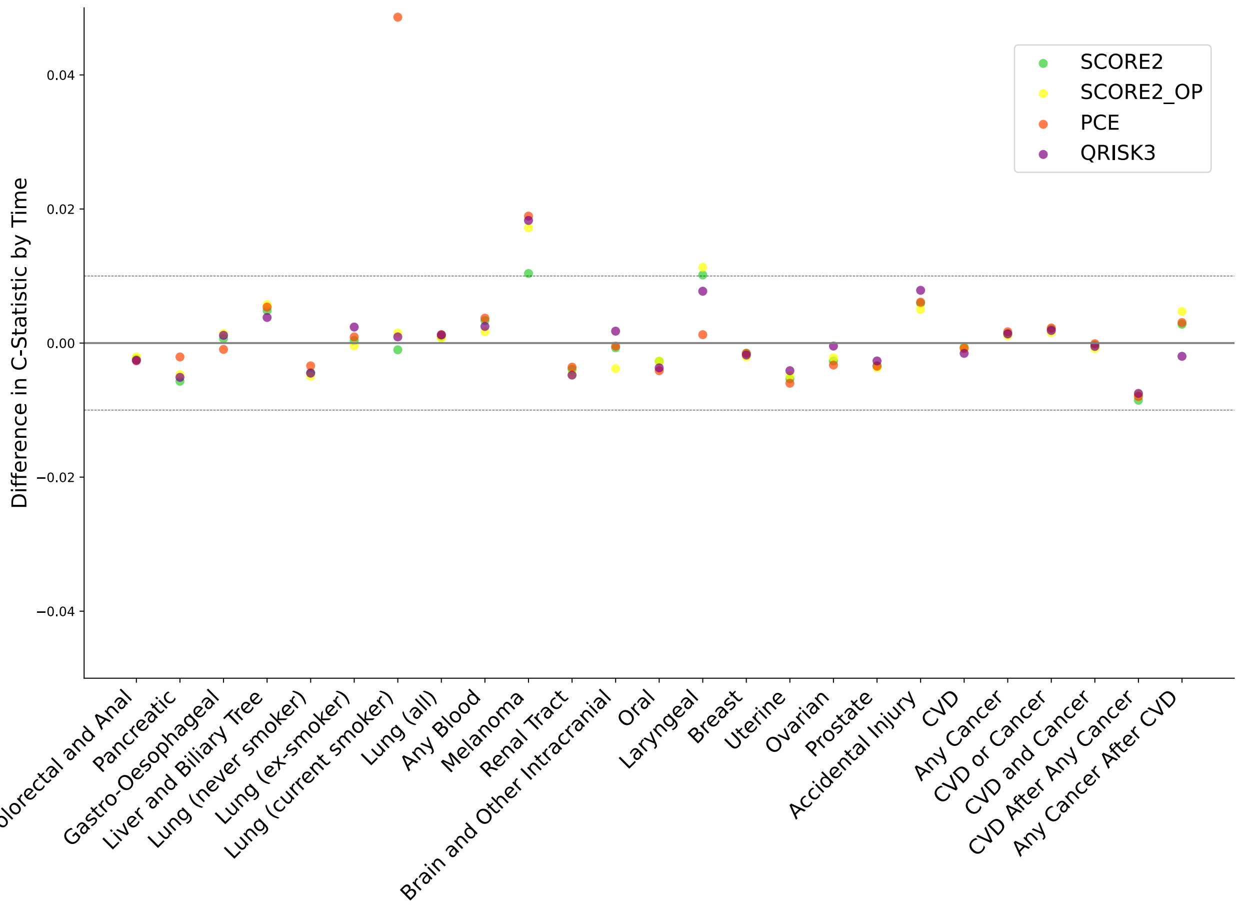

Figure S1: Miami plot showing the difference in c-statistic by follow-up time for the four CVD risk prediction models (QRISK3, PCE, SCORE2 and SCORE2-OP) for the various cancer outcomes. Most conditions show minimal variation in c-statistics, indicating predictions were stable over the different follow-up times.

**Figure S2: Significance of Time-Related Changes in Prediction Performance**

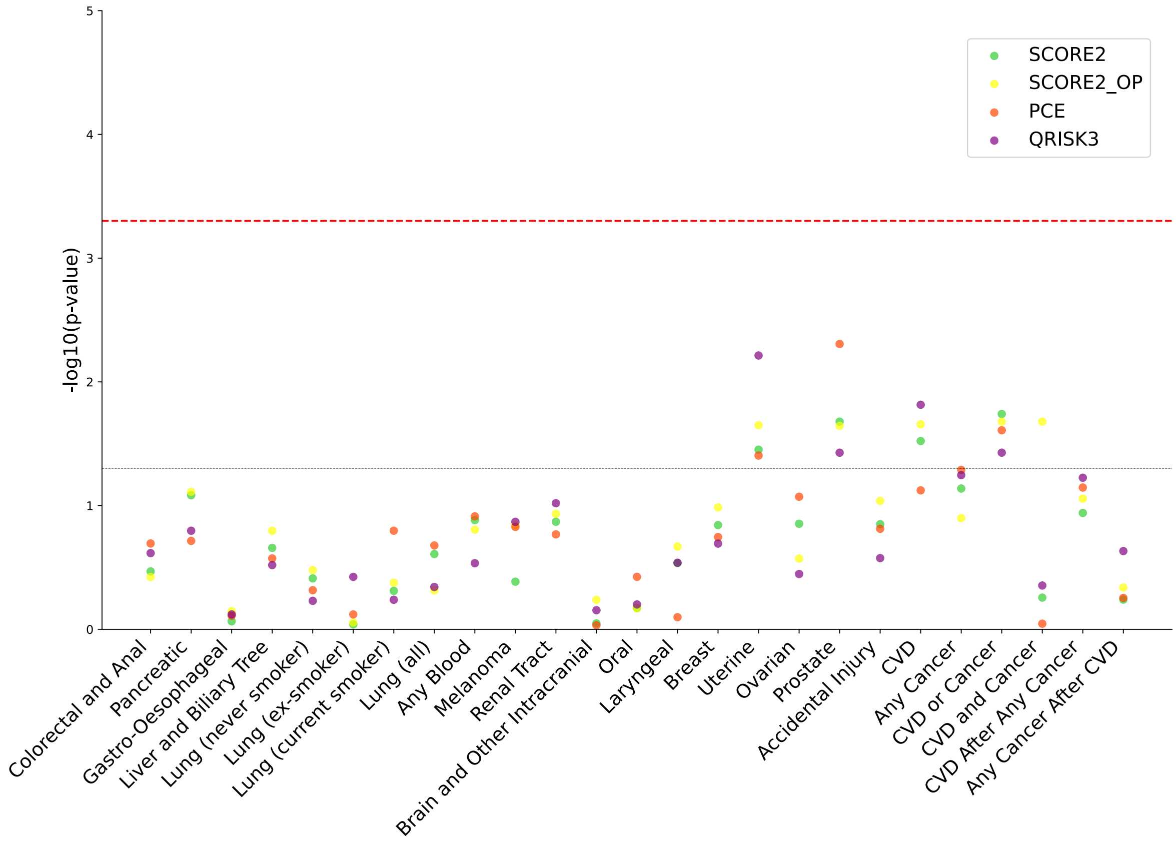

Figure S2: Manhattan plot showing the statistical significance (-log10(p values)) of the effect of follow-up time on c-statistic for each model (QRISK3, PCE, SCORE2, and SCORE2-OP) and outcome. The red dashed line indicates the threshold for statistical significance after Bonferroni correction.

**Figure S3. A comparison of the discrimination of each model between the UK Biobank and CPRD datasets.**

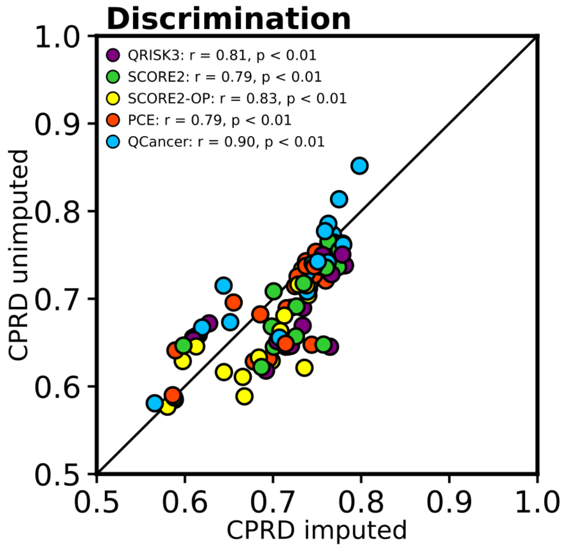

Figure S3: A comparison between model discrimination in UKB and the imputed CPRD dataset. The correlation (r) was calculated using Spearman's rank correlation test. The diagonal line indicates perfect correlation. The colours of the dots represent to each of the models evaluated (see legend). Participants with missing predictor or outcome data were excluded from both cohorts. For each outcome, those with prevalent disease were also excluded (see Supplementary Table S4-S5). Abbreviations: CPRD, Clinical Practice Research Datalink; UKB, UK Biobank.

**Figure S4: flow chart of patient selection in the UKB for predicting the any cancer outcome.**

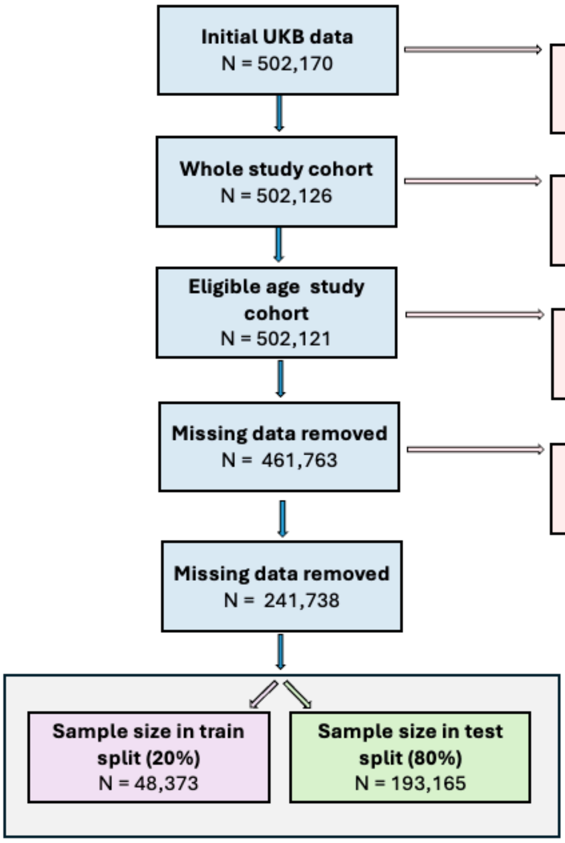

Figure S4: Flow chart summarising the number of participants excluded while preparing the UK biobank cohort to investigate model performance on predicting onset of any cancer.

**Figure S5: flow chart of patient selection in the complete case analysis of CPRD and imputed CPRD dataset for predicting the any cancer outcome.**

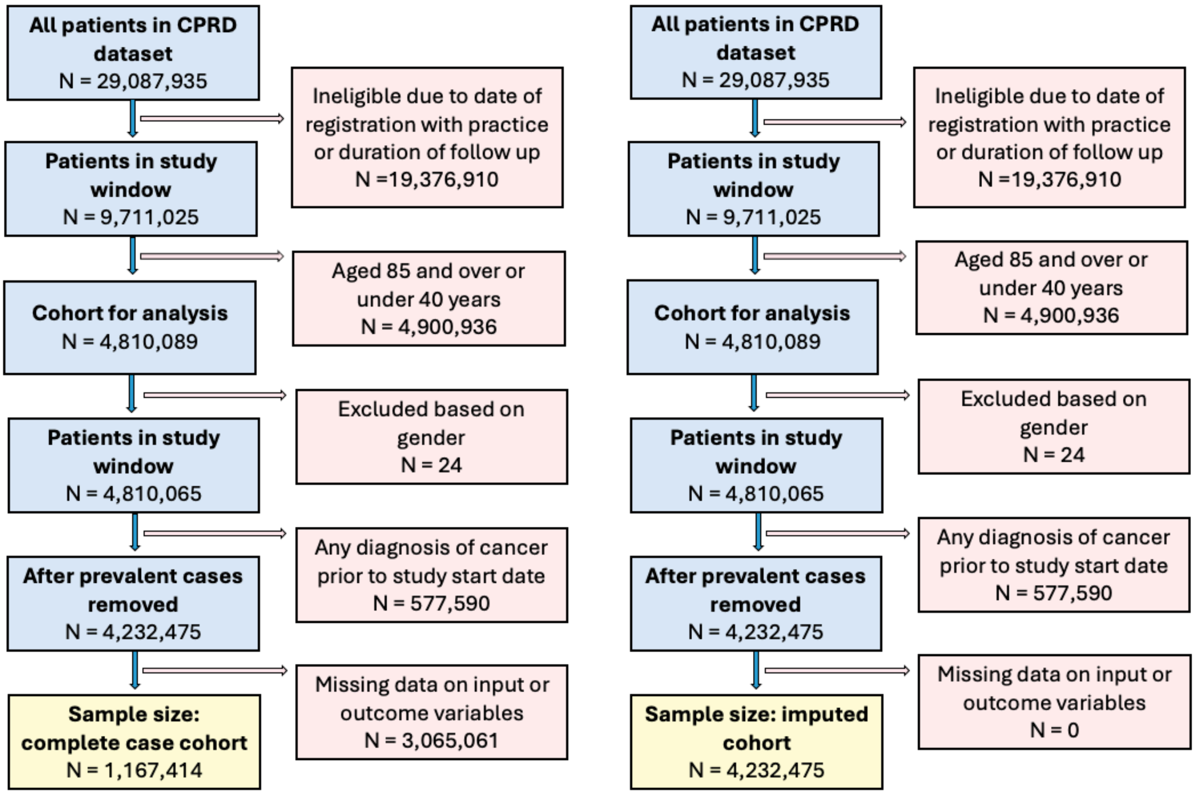

Figure S5: Flow chart summarising the number of participants excluded while preparing the CPRD complete case (left) and imputed (right) cohort to investigate model performance on predicting onset of any cancer.

**Figure S6: Difference in frequency of cancer types occurring before and after a CVD diagnosis.**

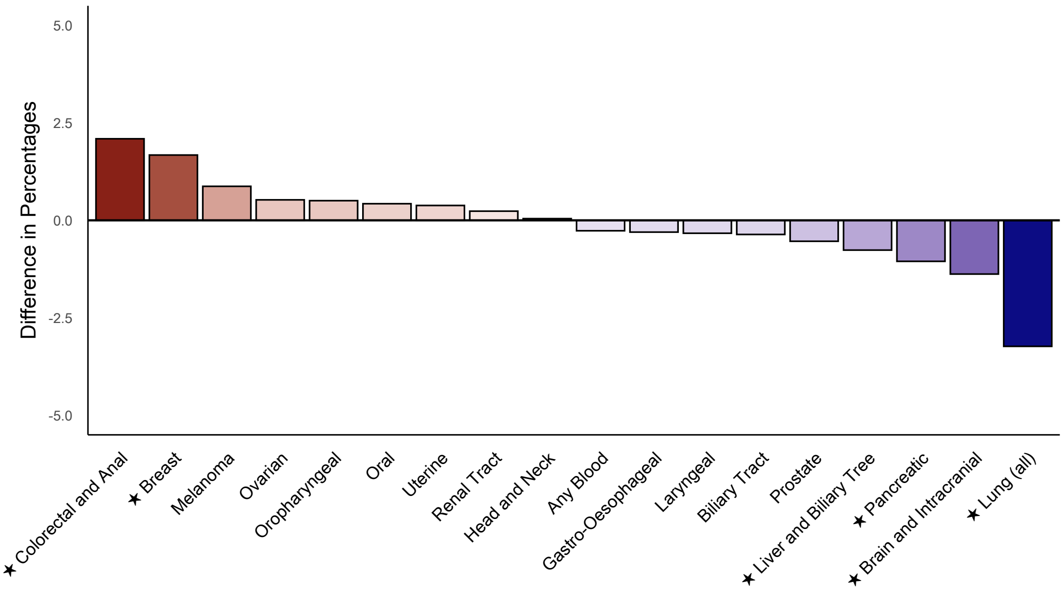

Figure S6: Bar chart showing the percentage difference between cancer diagnoses occurring before versus after cardiovascular disease (CVD) in participants who developed both (3,954 (1·00%) participants of the 203,831 of the UKB cohort without a prior diagnosis of CVD or cancer, and who were not taking lipid lowering medications at recruitment). Positive values (in red) refer to cancer types more frequently diagnosed before CVD and negative values (in blue) refer to cancers more commonly diagnosed after CVD. Cancer types with stars beside their names on the y-axis indicate statistically significant differences (p<0·05 after Bonferroni correction).

**Figure S7: Time until 10%, 15% and 50% mortality for difference cancers compared to their frequency of occurring before or after CVD in people who develop both.**

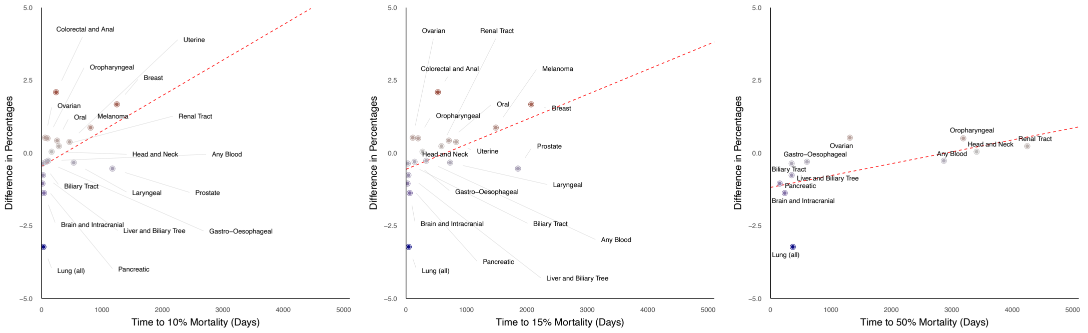

Figure S7 compares the typical sequence of a of cancer diagnosis relative to CVD in someone who developed both conditions with the survival times for a person diagnosed with each cancer. The y-axis represents the relative frequency of cancer occurring before (positive values) versus after (negative values) CVD diagnosis in participants who developed both conditions. The x-axis shows the time (in years) to reach 10% mortality (first panel), 15% mortality (second panel), and 50% mortality (third panel) for each cancer type. The number of participants who developed each cancer in the whole UKB cohort (n=502,121) can be found in Supplementary Table S4.

**Figure S8: Incidence of cancer cases by years after entering study start date.**

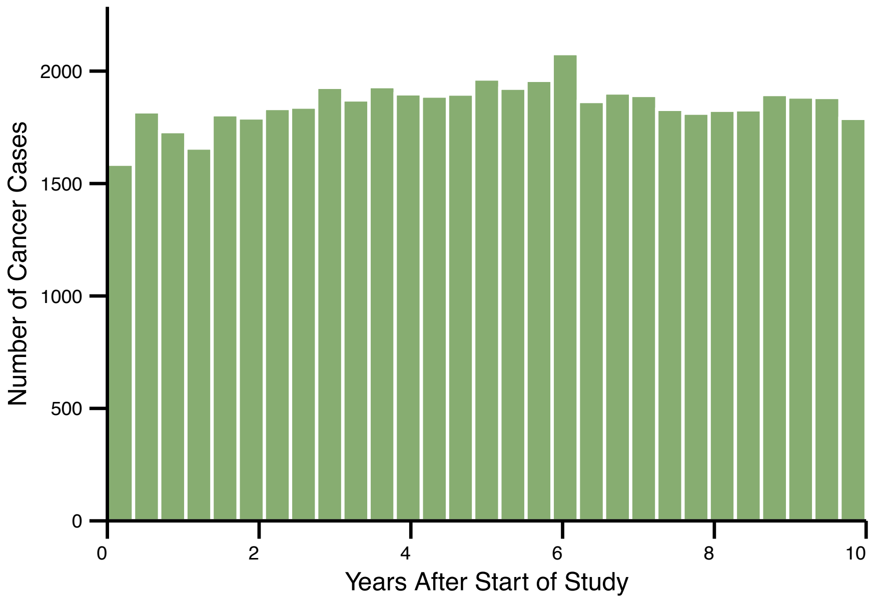

Figure S8: Bar chart showing the number of incident cancer cases diagnosed in UKB participants by years after study start date. The total number of participants was 502,121 (after excluding 5 participants who were outside of the target age range of under 40 or 85 and older). The total number of incident cancers over 10 years of follow-up was 53,821 (11·7%).
